# Impact of acute hospitalisation on development of long-term disease and health inequality: a longitudinal population study

**DOI:** 10.64898/2026.04.25.26351727

**Authors:** Yize I Wan, Rupert M Pearse, John R Prowle

**Author notes:** Correspondence to Yize I Wan PhD, Adult Critical Care Unit, The Royal London Hospital, Whitechapel, London E1 1FR.

## Abstract

**Objective:** To examine the impact of acute illness on long-term health and describe any differences in these associations between socioeconomic and ethnic groups.

**Design:** Longitudinal population study.

**Setting:** Linked primary and secondary care data recorded in the Clinical Practice Research Datalink (CPRD).

**Participants:** Adults (≥18 years) residing in England registered with a primary care general practice (GP) between 1st January 2012 and 31st December 2022 who have not opted out of inclusion into CPRD and linked data sources. Socioeconomic deprivation was defined using the Index of Multiple Deprivation (IMD) and ethnicity by UK census 2011 definitions.

**Main outcome measures:** The primary outcome was new long-term disease and multimorbidity (two or more long-term diseases). We describe incidence of hospitalisation for acute illness as the exposure.

**Results:** We included 18,329,659 people, with 9,339,394 (51.0%) women, 7,430,555 (40.5%) people from the most deprived deciles (IMD 1-4) and 3,009,717 (16.4%) from a minority ethnic group. 6,038,272 (32.9%) people experienced hospitalisation for acute illness. Hospitalisation was associated with increased onset of long-term disease in those alive at the end of follow up (41.1% hospitalised vs 18.7% not hospitalised; adjusted HR 2.48 (2.47 to 2.48)). Compared to non-hospitalised, those who had been hospitalised were more likely to change from being disease free at baseline to having a new long-term disease (12.9% vs. 7.5%), develop multimorbidity (4.7% vs. 1.1%), or transition to multimorbidity if they had pre-existing disease (8.1% vs. 1.8%). Age-standardised hospitalisation rates were highest in the most deprived decile and in people with Black ethnicity. Comparative hospitalisation ratio for IMD 1 compared to IMD 10 ranging from 1.78 in 2018 to 1.96 in 2021 and for Black ethnicity compared to White ranging from 1.03 in 2017 to 1.08 in 2021.

**Conclusions:** Acute hospitalisation is a key stage in the development of long-term disease and may be an underutilised opportunity for intervention to change healthy life trajectory and reduce health inequality.

**Summary box:** *What is already known on this topic:*

- Whilst inequalities in rates of long-term disease are well described, there is little evidence to describing the presence or absence of inequalities in significant acute illness and acute hospitalisation.
- The consequences of acute hospitalisation for acute illness on development of long-term disease are not well understood.

*What this study adds:*

- Acute hospitalisation is strongly associated with subsequent risk of developing long-term disease and multimorbidity.
- Age-standardised hospitalisation rates increased with higher levels of deprivation and was highest for people with Black ethnicity.
- People from socioeconomically deprived backgrounds and minority ethnic groups experience a reduced healthy life expectancy following acute hospitalisation.

*How this study might affect research, practice, or policy:*

- Acute hospitalisation may be an important marker for inequalities in healthy life expectancy and could be a key opportunity to better manage long-term health to reduce further inequalities.

## Introduction

While there have been improvements in healthcare provision and outcomes for the overall UK population, this has been accompanied by worsening health inequalities.^1^ Acute illness, like long-term health, can be a consequence of health inequality.^2^ This was clearly demonstrated by the COVID-19 pandemic.^3^ Inequalities including poor access to primary care services, and preventative healthcare, leave some people under-served by health systems increasing the risk of significant acute illness requiring hospitalisation.^4^ Significant acute illness often represents the start of new long-term disease, e.g. myocardial infarction leading to heart failure but may also represent an opportunity to recognise pre-existing undiagnosed long-term disease. Disadvantaged communities may be more likely to present to secondary care at a point of acute decline in baseline health compared to less acute interactions with primary care. Understanding determinants of health inequalities in acute illness may therefore be important in reducing inequalities in long-term health.

Our previous research has shown that in east London, acute hospitalisation is more frequent and occurs at a younger age among socioeconomically deprived and minority ethnic groups.^5-7^ These population groups also experience a higher prevalence of long-term disease - a major determinant of poor health outcomes. We do not know if this regional pattern extends more widely across the UK or exists in other nations. The long-term consequences of acute illness are not well understood. Multiple events can occur during acute hospitalisation and deteriorations not obviously related to the presenting illness are often not identified or followed up e.g. cardiac dysfunction after non-cardiac surgery or pneumonia complicated by acute kidney injury which often progresses to chronic kidney disease.^8,9^ This is not widely recognised as a cause of long-term poor health.^9^ Because long-term disease is itself a risk factor for hospitalisation with acute illness, patients may experience a cyclical decline in long-term health.

Acute hospitalisation may represent both a key indicator of worsening health and an opportunity for early intervention to prevent health inequalities. Contact with healthcare during acute illness may allow both recognition and management of previously undiagnosed long-term diseases as well as identify opportunities for primary and secondary prevention of new long-term diseases. We conducted an epidemiological study using linked routine primary and secondary care data in England to examine the impact of acute illness on long-term health and describe any differences in these associations between socioeconomic and ethnic groups. We aimed to test the hypothesis that acute hospitalisation is associated with development of long-term disease.

## Methods

### Data source

We followed REporting of studies Conducted using Observational Routinely-collected Data (RECORD) guidelines (supplement). We used anonymised, routinely collected electronic health record data from the Clinical Practice Research Datalink (CPRD) database and linked datasets covering around 25% of the UK general population representative in terms of age, sex, and ethnicity.^10^ We included data from both CPRD GOLD and CPRD Aurum containing primary care data collected using Vision® or EMIS® software systems respectively. Linked data to National Health Service hospitals in England was available from Hospital Episode Statistics (HES) up to 31st March 2021 including information on emergency department attendances using HES Accident and Emergency (HES A&E), hospitalisations using HES Admitted Patient Care (HES APC), and outpatient care (HES OP).^11^ Linked data to HES maternity were used to identify hospitalisation related to maternity care. Patient postcode linked socioeconomic deprivation measures were mapped using the composite 2019 English Index of Multiple Deprivation (IMD). Linkage to death registration data from the Office for National Statistics (ONS) was obtained. Postcode linked IMD was missing for 0.1% of the cohort. Linkage to all other datasets were complete. Access to data was approved via the CPRD research data governance process and this study was conducted in accordance with a pre-specified protocol (CPRD study protocol: 21_000720).

### Study cohort

We included all adults (≥18 years) residing in England registered with a primary care general practice (GP) between 1st January 2012 and 31st December 2022 who have not opted out of inclusion into CPRD and linked data sources. All individuals had at least 365 days of follow-up from date of GP registration and were followed up until the earliest of date of last data collection, transfer out of practice, or death.

### Definitions of key variables

The exposure was acute hospitalisation during the follow-up period. Prior admissions outside of this window were not considered due to incomplete historical data recording of hospitalisation. Hospitalisations related to maternity care and elective admissions including waiting list, booked, and planned admissions e.g., for surgery, chemotherapy, or dialysis were excluded. Accordingly, acute hospitalisation represents a hospital admission for investigation and/or treatment of an acute illness, either as a new condition or exacerbation of a pre-existing one. The first hospitalisation during the follow-up period was identified as the index hospitalisation. Individual-level relative measures of socioeconomic deprivation were assessed using IMD deciles (1st, most deprived). People with missing IMD were excluded from comparative analyses. Ethnicity was defined using the CPRD Ethnicity Record dataset (Asian, Black, Mixed/multiple, White, Other, Unknown) corresponding to UK Census 2011 higher-level categorisations.^12^ People with unknown ethnicity were included in comparative analyses as misclassification of non-White individuals into unknown or other ethnic groups occurs more frequently in emergency admissions. We examined age, sex, smoking history, alcohol intake, and obesity as additional baseline variables.

### Outcome measures

The primary outcome was development of new long-term disease and multimorbidity (two or more long-term diseases). We assessed all individuals including those with long-term disease at baseline as we aimed to examine acquisition of new long-term disease. We focused on long-term diseases included in the Charlson Comorbidity Index and previous multimorbidity studies.^13,14^ Diagnoses were identified from primary care records using phenotyping algorithms available online (full definitions in Table S1). Secondary outcomes were age at development of first long-term disease and age at development of multimorbidity at any point including preceding the follow-up period, and in those who experienced hospitalisation during the follow-up period we report: age at first hospitalisation, total numbers of hospitalisations, multiple hospitalisations, and change in healthcare contact days comparing the year before and after acute hospitalisation.

### Statistical analysis

We compared baseline characteristics between those who had experienced at least one hospitalisation and those who had not been hospitalised in the study period. Our first objective was to examine rates of hospitalisation based on IMD decile and ethnicity category. To account for potential differences in age structure between IMD deciles and between ethnic groups, we calculated comparative hospitalisation ratios using age-standardised hospitalisation rates based on ONS England population data for each year during the study period. Our second objective was to examine the association between acute hospitalisation on the acquisition of new long-term disease including those being diagnosed with their first long-term disease and those with pre-existing disease who are developing new or worsening multimorbidity. Cox proportional hazards models censored for death were used to assess the relationship between acute hospitalisation as a time-dependent covariate and development of subsequent long-term disease adjusted for age, sex, and baseline number of long-term diseases (categorised as none, one to two, more than two). To account for non-linear effects of age, a restricted cubic spline model with five knots was used. People with unknown sex were excluded. Data are presented as mean (SD), median (IQR) or n (%). Effect measures are presented as hazards ratios (HR) with 95% confidence intervals (CI). All analyses were performed using RStudio 2025.05.1-513 and R version 4.5.1 (Rocky9) using OnDemand.^15^

### Sensitivity analysis

As a sensitivity analysis, we repeated the Cox proportional hazards analysis including death as a combined outcome measure.

### Exploratory analysis

We carried out an exploratory analysis to describe differences in age of first hospitalisation between IMD deciles and ethnicity groups considering differences in age distribution between IMD and ethnicity subgroups.

### Patient and public involvement

This study involved secondary use of electronic healthcare records collected as part of routine clinical practice. We consulted our panel of patient and public representatives in the reporting and interpretation of study findings and implications for future work.

## Results

We included a total of 18,329,659 individuals (Fig. S1). Baseline characteristics for patients were similar between CPRD GOLD and Aurum datasets (Table S2, Figs. S2 and S3). Between 2012 and 2021, 6,038,272 (32.9%) had experienced an acute hospitalisation. Baseline characteristics for the total cohort and stratified by hospitalisation are presented in Table 1. Indications for hospitalisation grouped by admitting specialty are listed in Table S3. People who had been hospitalised were older and a greater proportion were female. Hospitalised individuals were more likely to have a history of smoking, moderate to heavy alcohol use, obesity, and all long-term diseases in particular cardiovascular conditions (heart failure, coronary heart disease, cerebrovascular disease), dementia, and peripheral vascular disease.

**Table 1.**
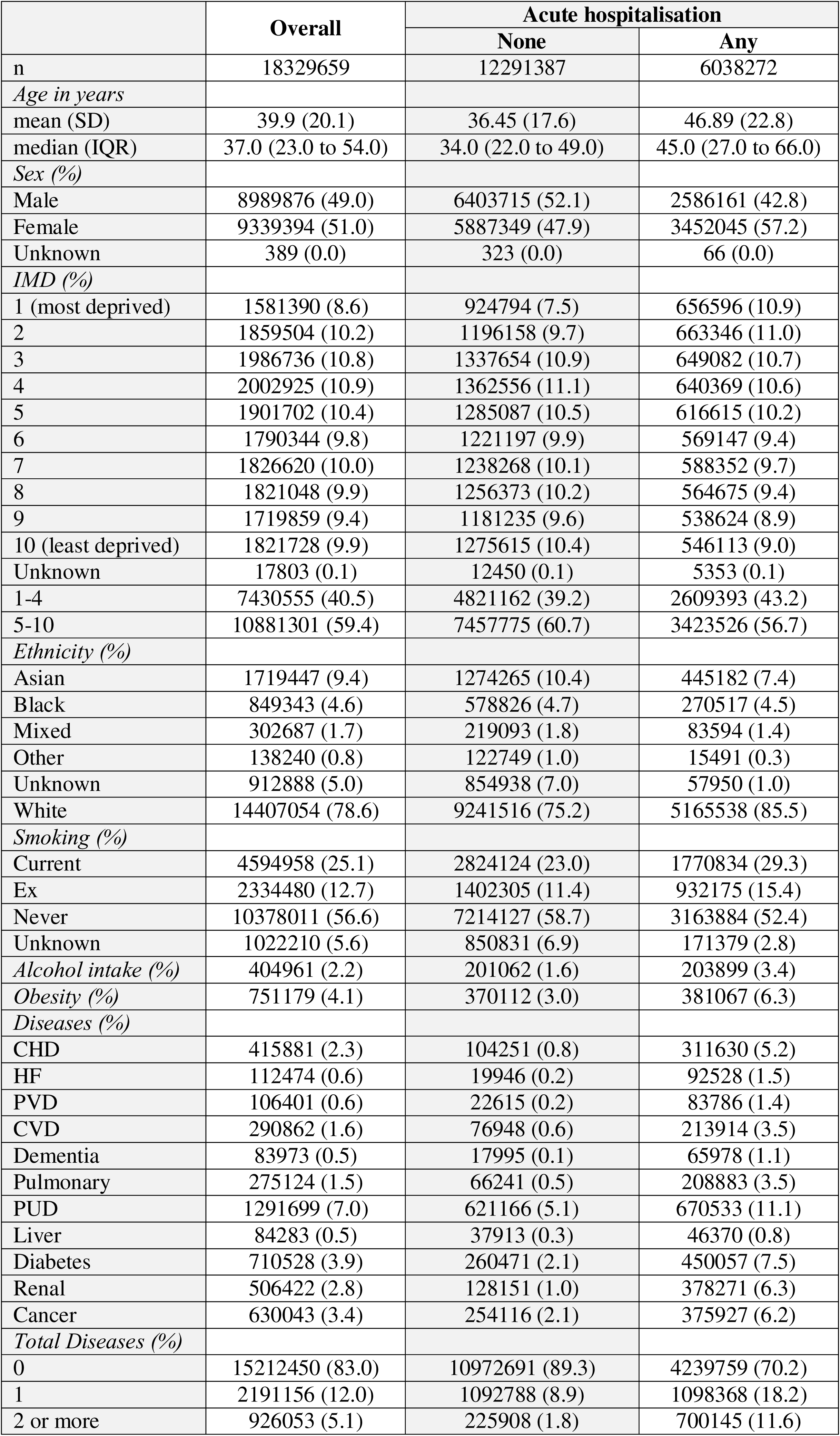
Baseline characteristics at start of follow-up 01/01/2012, stratified according to acute hospitalisation. All data presented as n (%) unless otherwise indicated, proportion by rows. IMD: index of multiple deprivation, CHD: coronary heart disease, HF: heart failure, PVD: peripheral vascular disease, PUD: peptic ulcer disease, CVD: cerebrovascular disease.

|  | Overall | Acute hospitalisation |  |
| --- | --- | --- | --- |
|  |  | None | Any |
| n | 18329659 | 12291387 | 6038272 |
| *Age in years* |  |  |  |
| mean (SD) | 39.9 (20.1) | 36.45 (17.6) | 46.89 (22.8) |
| median (IQR) | 37.0 (23.0 to 54.0) | 34.0 (22.0 to 49.0) | 45.0 (27.0 to 66.0) |
| *Sex (%)* |  |  |  |
| Male | 8989876 (49.0) | 6403715 (52.1) | 2586161 (42.8) |
| Female | 9339394 (51.0) | 5887349 (47.9) | 3452045 (57.2) |
| Unknown | 389 (0.0) | 323 (0.0) | 66 (0.0) |
| *IMD (%)* |  |  |  |
| 1 (most deprived) | 1581390 (8.6) | 924794 (7.5) | 656596 (10.9) |
| 2 | 1859504 (10.2) | 1196158 (9.7) | 663346 (11.0) |
| 3 | 1986736 (10.8) | 1337654 (10.9) | 649082 (10.7) |
| 4 | 2002925 (10.9) | 1362556 (11.1) | 640369 (10.6) |
| 5 | 1901702 (10.4) | 1285087 (10.5) | 616615 (10.2) |
| 6 | 1790344 (9.8) | 1221197 (9.9) | 569147 (9.4) |
| 7 | 1826620 (10.0) | 1238268 (10.1) | 588352 (9.7) |
| 8 | 1821048 (9.9) | 1256373 (10.2) | 564675 (9.4) |
| 9 | 1719859 (9.4) | 1181235 (9.6) | 538624 (8.9) |
| 10 (least deprived) | 1821728 (9.9) | 1275615 (10.4) | 546113 (9.0) |
| Unknown | 17803 (0.1) | 12450 (0.1) | 5353 (0.1) |
| 1-4 | 7430555 (40.5) | 4821162 (39.2) | 2609393 (43.2) |
| 5-10 | 10881301 (59.4) | 7457775 (60.7) | 3423526 (56.7) |
| *Ethnicity (%)* |  |  |  |
| Asian | 1719447 (9.4) | 1274265 (10.4) | 445182 (7.4) |
| Black | 849343 (4.6) | 578826 (4.7) | 270517 (4.5) |
| Mixed | 302687 (1.7) | 219093 (1.8) | 83594 (1.4) |
| Other | 138240 (0.8) | 122749 (1.0) | 15491 (0.3) |
| Unknown | 912888 (5.0) | 854938 (7.0) | 57950 (1.0) |
| White | 14407054 (78.6) | 9241516 (75.2) | 5165538 (85.5) |
| *Smoking (%)* |  |  |  |
| Current | 4594958 (25.1) | 2824124 (23.0) | 1770834 (29.3) |
| Ex | 2334480 (12.7) | 1402305 (11.4) | 932175 (15.4) |
| Never | 10378011 (56.6) | 7214127 (58.7) | 3163884 (52.4) |
| Unknown | 1022210 (5.6) | 850831 (6.9) | 171379 (2.8) |
| *Alcohol intake (%)* | 404961 (2.2) | 201062 (1.6) | 203899 (3.4) |
| *Obesity (%)* | 751179 (4.1) | 370112 (3.0) | 381067 (6.3) |
| *Diseases (%)* |  |  |  |
| CHD | 415881 (2.3) | 104251 (0.8) | 311630 (5.2) |
| HF | 112474 (0.6) | 19946 (0.2) | 92528 (1.5) |
| PVD | 106401 (0.6) | 22615 (0.2) | 83786 (1.4) |
| CVD | 290862 (1.6) | 76948 (0.6) | 213914 (3.5) |
| Dementia | 83973 (0.5) | 17995 (0.1) | 65978 (1.1) |
| Pulmonary | 275124 (1.5) | 66241 (0.5) | 208883 (3.5) |
| PUD | 1291699 (7.0) | 621166 (5.1) | 670533 (11.1) |
| Liver | 84283 (0.5) | 37913 (0.3) | 46370 (0.8) |
| Diabetes | 710528 (3.9) | 260471 (2.1) | 450057 (7.5) |
| Renal | 506422 (2.8) | 128151 (1.0) | 378271 (6.3) |
| Cancer | 630043 (3.4) | 254116 (2.1) | 375927 (6.2) |
| *Total Diseases (%)* |  |  |  |
| 0 | 15212450 (83.0) | 10972691 (89.3) | 4239759 (70.2) |
| 1 | 2191156 (12.0) | 1092788 (8.9) | 1098368 (18.2) |
| 2 or more | 926053 (5.1) | 225908 (1.8) | 700145 (11.6) |

### Age-standardised hospitalisation by IMD and ethnicity

Within the cohort, 7,430,555 (40.5%) of people resided in the most deprived four IMD deciles 1-4. The ethnicity distribution was majority White (n=14,407,054, 78.6%), followed by Asian (n=1,719,447, 9.4%), Black (n=849,343, 4.6%), Mixed (n=302,687, 1.7%), and Other (n=138,240, 0.8%). 5% of the cohort were of unknown ethnicity (n=912,888) (Table 1). People from minority ethnic groups were more deprived (Fig. S4). Overall rates of hospitalisation increased every year during the follow-up period. Age-standardised hospitalisation rates increased with increasing deprivation: comparative hospitalisation ratio for IMD 1 compared to IMD 10 ranging from 1.78 in 2018 to 1.96 in 2021 (Fig. 1, Fig. S5 and Table S4). There were also consistent differences in age-standardised hospitalisation rates across ethnic groups over time. Compared to White, people with Black ethnicity were more likely to be hospitalised with comparative hospitalisation rate ranging from 1.03 in 2017 to 1.08 in 2021. However, comparative hospitalisation ratios were lower for Asian, Mixed, Other and Unknown compared to White ethnicity (Fig. 2 and Table S4).

**Figure 1.**
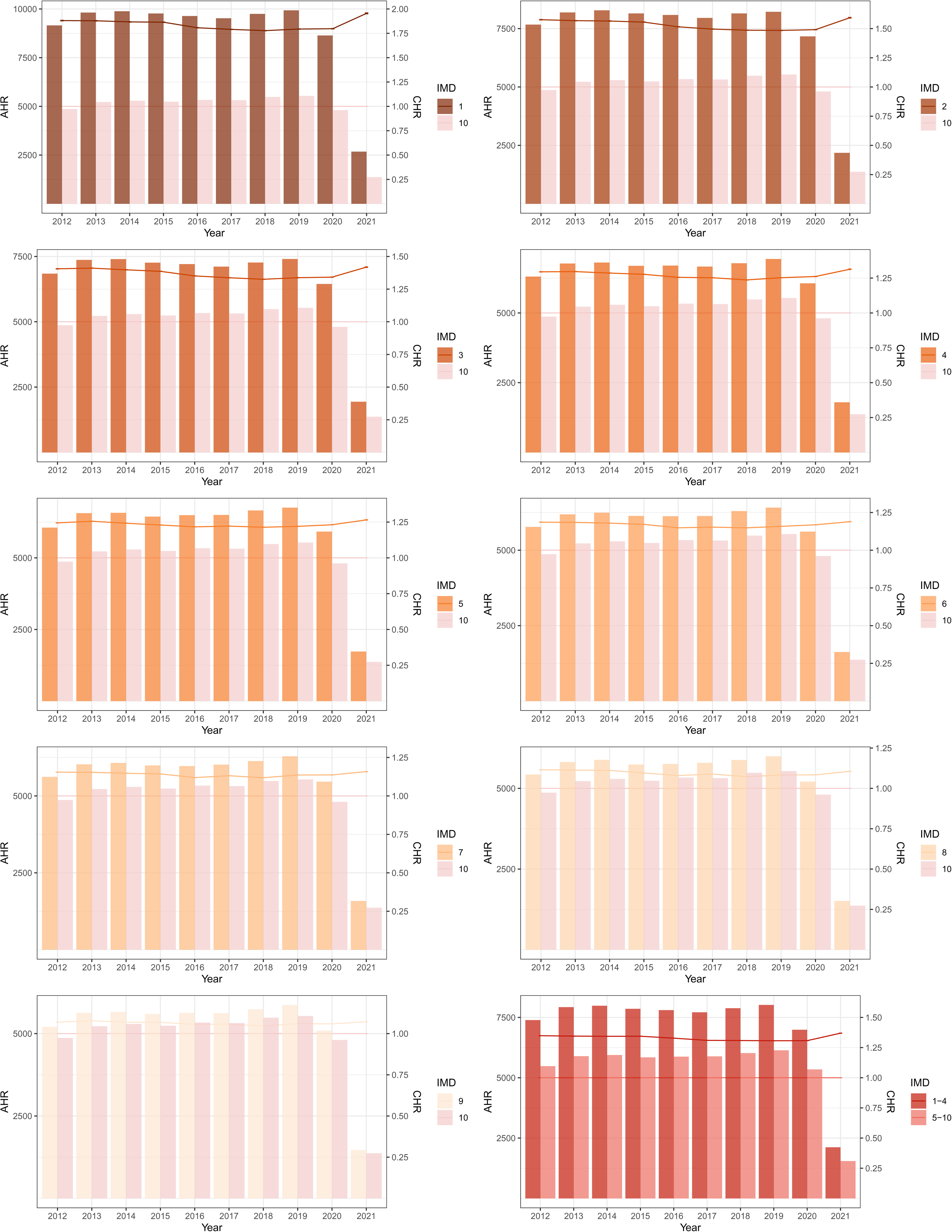
Age-standardised hospitalisation rate per 100,000 population per year and comparative hospitalisation ratio by IMD decile, plots by year. Bar charts showing the age-standardised hospitalisation rate (AHR) by Index of Multiple Deprivation decile (IMD 1 most deprived) using ONS population data per year, axis on the left-hand side of each plot. Line graph showing the comparative hospitalisation ratio (CHR) for each IMD decile compared to IMD 10 (least deprived), axis on the right-hand side of each plot. 95% CI shown by error bars. 2021 data to 31st March only.

**Figure 2.**
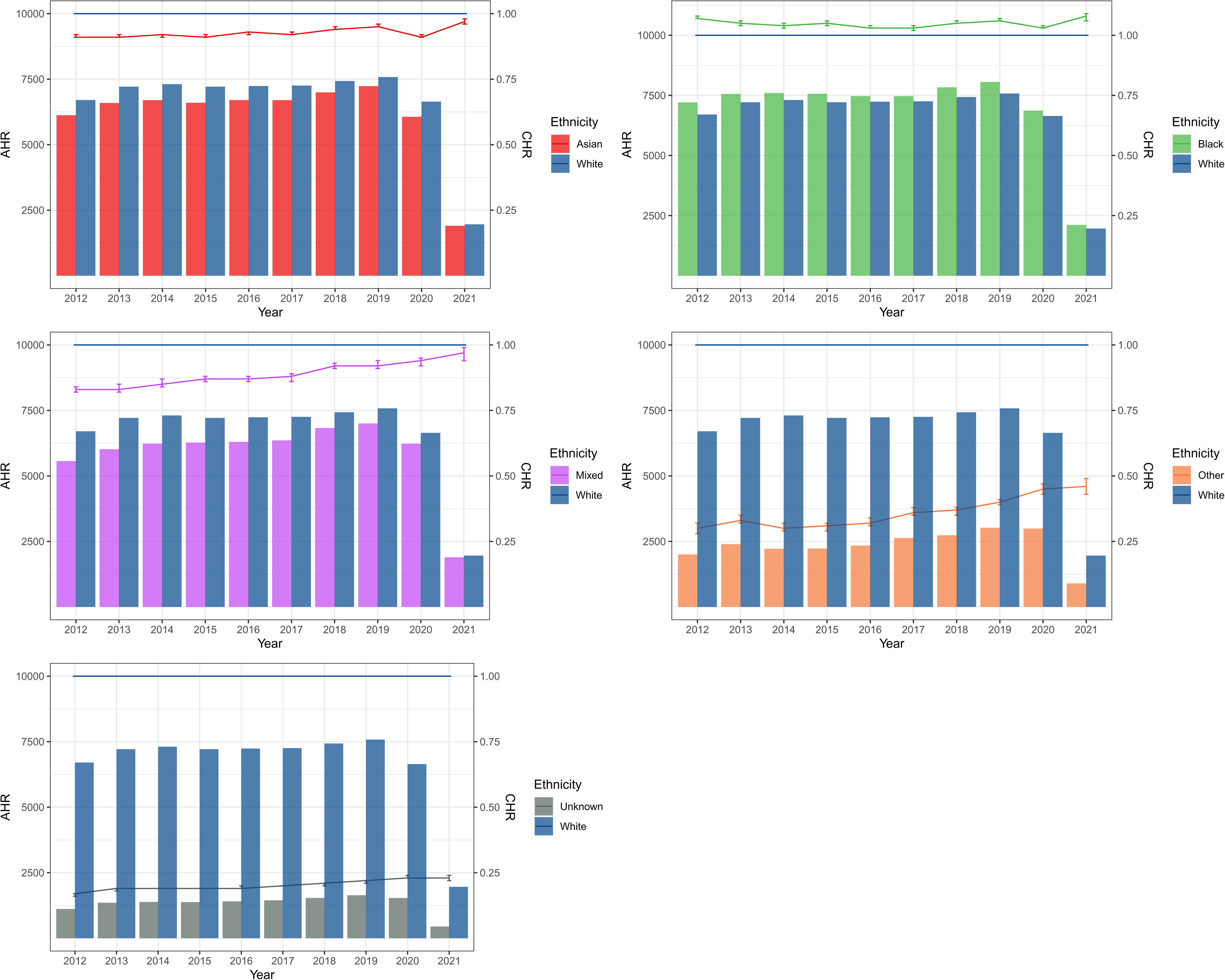
Age-standardised hospitalisation rate per 100,000 population per year and comparative hospitalisation ratio by ethnicity. Bar charts showing the age-standardised hospitalisation rate (AHR) per year using ONS population data by ethnicity, axis on the left-hand side of each plot. Line graph showing the comparative hospitalisation ratio (CHR) for each ethnicity compared to White, axis on the right-hand side of each plot. 95% CI shown by error bars. Categories compared in each plot are listed in the legend on the right of each plot. 2021 data to 31st March only.

### Development of long-term disease and multimorbidity

Overall, 3,261,464 (17.8%) people developed new long-term disease at the end of follow-up: 2,480,350 (13.6%) developed one and 772,114 (4.2%) developed multimorbidity (two or more diseases) (Table 2). Compared to people who had not been hospitalised, greater proportions of hospitalised people developed new long-term disease (31.6% vs. 10.9%). At the end of follow-up, 2,328,196 (22.0%) of hospitalised individuals had one long-term disease and 582,636 (9.6%) had multimorbidity compared to 1,161,154 (9.4%) and 189,478 (1.5%) respectively of non-hospitalised individuals. Compared to non-hospitalised, those who had been hospitalised were also more likely to change from being disease free at baseline to having a new long-term disease (12.9% vs. 7.5%), develop multimorbidity (4.7% vs. 1.1%), or transition to multimorbidity if they had pre-existing disease (8.1% vs. 1.8%) (Table 2). The diseases with highest prevalence were peptic ulcer disease, diabetes, and cancer in both groups. Hospitalised individuals had higher prevalence of all diseases in particular heart failure (10.7 times), dementia (8.3 times), and cerebrovascular disease (6.1 times) compared to non-hospitalised. Acute hospitalisation was associated with increased risk of long-term disease (HR 3.52, 95% CI 3.51 to 3.52). This attenuated to HR 2.48, 95% CI 2.47 to 2.48 after adjustment for age, sex, and baseline number of long-term diseases (Table S5). As a sensitivity analysis, including death as a combined outcome measure in the Cox model (instead on censoring at date of death) did not substantially alter the hazard ratio associated with hospitalisation but increased its absolute effect as baseline event rates was higher (Table S5 and Fig. S6). The absolute impact of acute hospitalisation varied for different patients, for example, an 80-year-old female with one-to-two long-term diseases at baseline was more likely to develop worsening multimorbidity compared to a 50-year-old male with two or more pre-existing diseases or a 30-year-old female with no pre-existing disease (Fig. 3).

**Figure 3.**
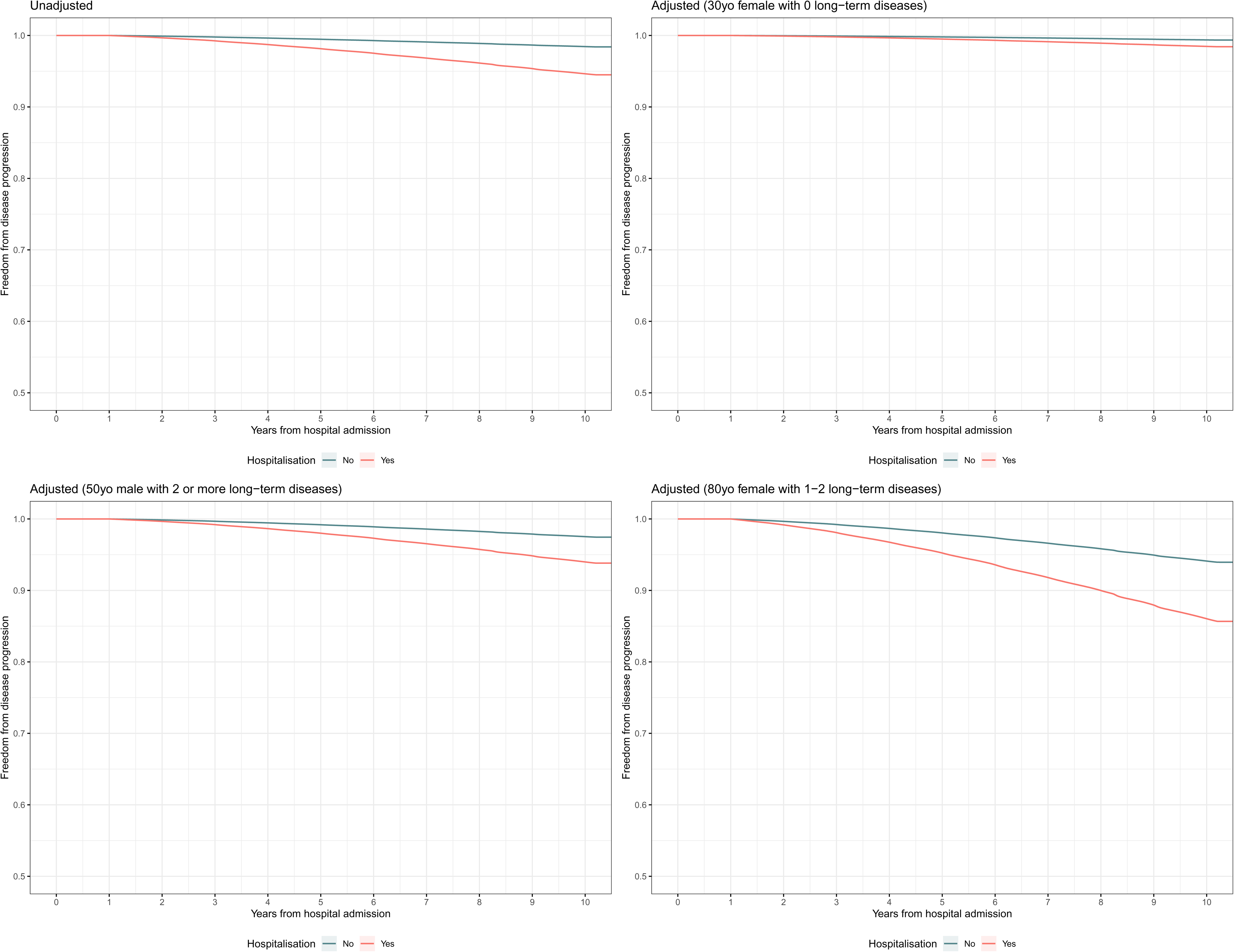
Time to event curves comparing acute hospitalisation. Adjusted for age, sex, and baseline number of long-term diseases. Y-axis censored at 0.50.

**Table 2.** Long-term disease diagnoses at start and end of follow up, stratified according to acute hospitalisation. All data presented as n (%) unless otherwise indicated, proportion by columns. CHD: coronary heart disease, HF: heart failure, PVD: peripheral vascular disease, PUD: peptic ulcer disease, CVD: cerebrovascular disease. *Proportion of those at start of follow-up.

|  | Overall |  | Acute hospitalisation |  |  |  |
| --- | --- | --- | --- | --- | --- | --- |
|  | Start | End | None |  | Any |  |
|  | Start | End | Start | End | Start | End |
| n | 18329659 | 17418353 | 12291387 | 12157311 | 6038272 | 5261042 |
| *Died (%)* * | - | 911306 (0.05) | - | 134076 (0.01) | - | 777230 (0.13) |
| *Number of new diseases (%)* * |  |  |  |  |  |  |
| 0 | - | 15068195 (82.2) | - | 10940755 (89.0) | - | 4127440 (68.4) |
| 1 | - | 2489350 (13.6) | - | 1161154 (9.4) | - | 1328196 (22.0) |
| 2 or more | - | 772114 (4.2) | - | 189478 (1.5) | - | 582636 (9.6) |
| *Change from baseline (%)* * |  |  |  |  |  |  |
| 0 to 1 | - | 1705077 (9.3) | - | 923635 (7.5) | - | 781442 (12.9) |
| 0 to 2 or more | - | 413599 (2.3) | - | 130679 (1.1) | - | 282920 (4.7) |
| 1 to 2 or more | - | 714117 (3.9) | - | 226679 (1.8) | - | 487438 (8.1) |
| *Total number (%)* |  |  |  |  |  |  |
| 0 | 15212450 (83.0) | 13093774 (86.1) | 10972691 (89.3) | 9880714 (81.3) | 4239759 (70.2) | 3098792 (58.9) |
| 1 | 2191156 (12.0) | 3182116 (17.4) | 1092788 (8.9) | 1749226 (14.4) | 1098368 (18.2) | 1196165 (22.7) |
| 2 or more | 926053 (5.1) | 2053769 (11.2) | 225908 (1.8) | 527371 (4.3) | 700145 (11.6) | 966085 (18.4) |
| *Diseases (%)* |  |  |  |  |  |  |
| CHD | 415881 (2.3) | 659626 (3.6) | 104251 (0.8) | 146476 (1.2) | 311630 (5.2) | 336455 (6.4) |
| HF | 112474 (0.6) | 351845 (1.9) | 19946 (0.2) | 41831 (0.3) | 92528 (1.5) | 168642 (3.2) |
| PVD | 106401 (0.6) | 195933 (1.1) | 22615 (0.2) | 39576 (0.3) | 83786 (1.4) | 87119 (1.7) |
| CVD | 290862 (1.6) | 564811 (3.1) | 76948 (0.6) | 110482 (0.9) | 213914 (3.5) | 286968 (5.5) |
| Dementia | 83973 (0.5) | 386495 (2.1) | 17995 (0.1) | 44967 (0.4) | 65978 (1.1) | 174600 (3.3) |
| Pulmonary | 275124 (1.5) | 548675 (3.0) | 66241 (0.5) | 140889 (1.2) | 208883 (3.5) | 249515 (4.7) |
| PUD | 1291699 (7.0) | 2301819 (12.6) | 621166 (5.1) | 1109091 (9.1) | 670533 (11.1) | 966189 (18.4) |
| Liver | 84283 (0.5) | 282006 (1.5) | 37913 (0.3) | 125617 (1.0) | 46370 (0.8) | 129763 (2.5) |
| Diabetes | 710528 (3.9) | 1268847 (2.6) | 260471 (2.1) | 507152 (4.2) | 450057 (7.5) | 554604 (10.5) |
| Renal | 506422 (2.8) | 910932 (5.0) | 128151 (1.0) | 237760 (2.0) | 378271 (6.3) | 402782 (7.7) |
| Cancer | 630043 (3.4) | 1302666 (7.1) | 254116 (2.1) | 484161 (4.0) | 375927 (6.2) | 478077 (9.1) |

### Secondary outcomes

In those who developed long-term disease, age at first long-term disease was 54 (40 to 66) years and age at multimorbidity was 69 (59 to 77) years (Tables S7 and S8). Similar differences in age were observed by level of deprivation and ethnicity with onset of both first long-term disease and multimorbidity occurring at a younger age in more deprived and minority ethnicity groups (Fig. S6). However, for people of Black ethnicity, the gradient across deprivation deciles was again very small (one year between most and least deprived) (Tables S7 and S8). Across the whole cohort, patients were on average hospitalised 0.9 times and 17.4% experienced two or more hospitalisations (Table S6). The average number and proportion of people with multiple hospitalisations increased with increasing deprivation (IMD 1: mean 1.4, 24.1% multiple vs. IMD 10: mean 0.7, 14.9% multiple). People from minority ethnic groups experienced fewer average numbers of hospitalisations and multiple hospitalisations (Table S6).

### Healthcare contact days

Comparing the year before and after acute hospitalisation, the total number of days alive spent in contact with a healthcare professional increased by a mean of 8.7 (SD 21.3) days (Fig. S7). Most of these days were spent in primary care. The magnitude of increase in contact days was slighter higher in less deprived patients (IMD 1: 8.3 (SD 20.8) vs. IMD 10: 9.0 (SD 21.5)) (Fig. S8). Higher proportions of contact days both before and after hospitalisation were being spent in primary care and outpatient appointments for less deprived compared to repeat A&E attendance and hospitalisation for more deprived patients. White and Black patients had the highest level of increase in contact days overall and in repeat hospitalisations (Fig. S9).

### Age at first hospitalisation

In this exploratory analysis, by the end of follow-up, 50% of those aged 70 years in IMD 1 had experienced a hospitalisation in the preceding eight years while in IMD 10 this proportion was not reached until age 80 years. There was a ten-year difference with a trend of increasing age across IMD (Table S7, Fig. S10). Age at first hospitalisation were comparable between ethnic groups at 77 years (White), 78 years (Asian), 79 years (Black) and 80 years (Mixed) (Table S7, Fig. S10). People with other ethnicity were excluded from this analysis as this group had substantially fewer proportions of hospitalisation across the age range with <50% throughout.

## Discussion

The principal finding was that people from deprived IMD deciles and certain minority ethnic groups had higher rates of hospital admission than expected for their age profile. Importantly, acute hospitalisation increased subsequent risk of developing long-term disease by over two times. This suggests that people who are socioeconomically deprived and from minority ethnic backgrounds experience a shorter healthy life expectancy and that inequalities in long-term health are at least in part driven by inequalities in acute illness. We also demonstrate variations in level of socioeconomic deprivation across ethnicities. These are to the best of our knowledge the first and largest published UK data to report on acute healthcare use across socioeconomic and ethnic groups.

### Comparison to previous studies

There are limited studies examining inequalities in acute illness and acute healthcare use. The majority predate 2013, are based on single centre or regional datasets, self-reported data, and/or focus on subgroups of the acutely ill population meaning findings may no longer be representative or widely generalisable. These studies also report higher incidence of illness and worsening outcomes with increased deprivation and minority ethnic background. Two more recent studies examining critical care outcomes in Scotland and patients with severe sepsis in the US report higher short-term mortality rates in more deprived patients.^16,17^ Previous studies examining ethnicity were frequently limited by lack of comprehensive adjustment for other risk factors including socioeconomic status and long-term diseases. We found that the influence of socioeconomic deprivation varied across ethnic groups such that age at first hospitalisation did not change by level of deprivation in Black patients. This suggests that disadvantage in Black patients was more related to their ethnicity and that even the least deprived Black patients were as disadvantaged as the most deprived White patients in terms of associations of acute illness. This supports previous studies showing different effects of socioeconomic determinants within different ethnicities.^18,19^

### Quantifying health inequalities

NHS data comparing healthcare use by index of deprivation and ethnic categories are variably reported across nations and were only reported for the first time in 2018. Cross-sectional summary data from routine electronic health records in England and Scotland have shown that people from more deprived socioeconomic backgrounds and minority ethnic groups have higher rates of emergency A&E attendance.^20,21^ In England, mean ages at hospitalisation are also younger in these patient populations.^22^ Our findings support these statistics and provide longitudinal individual-level data linked across primary and secondary care. This study supports our previous work showing that regional patterns seen in east London extend more widely across England. Importantly, it highlights the fact that some differences in healthcare outcomes may be masked by statistical adjustment using measures such as mortality as patients from deprived and minority ethnic backgrounds have lower rates of mortality following hospitalisation due to being admitted at a younger age. When a person arrives in hospital and is admitted acutely for the first time in their life, on average, patients from more deprived background or some minority groups will be younger. This is likely to represent a mix of getting acute conditions at a younger age, less ability to manage such conditions without admission, and an overall larger proportion of younger people in more deprived areas. All of these are important health inequalities that may replicate to development of long-term disease and underline the importance of the acute admission as an opportunity for both acute treatment and long-term disease prevention and management particularly in more deprived populations.

### Long-term consequences of acute illness

Physical and functional decline in the elderly following acute hospitalisation has been clearly documented and led to multiple multidisciplinary and therapy-based interventions.^23^ There is also increasing evidence for persistent deficits and healthcare utilisation following severe and critical illness.^24,25^ Few studies have examined the long-term health consequences of acute illness more generally including among younger adults without pre-existing disease. Acute hospital admissions are often poorly specified, and presentations often do not reflect a single illness or disease process. As such, we have examined hospital contact through secondary care both as an exposure and an opportunity for intervention to address long-term health trajectory. We have shown that acute illness and hospitalisation have a significant impact in worsening long-term health through the development of long-term disease and multimorbidity. The most prevalent resulting long-term diseases such as cardiovascular disease, cerebrovascular disease, and dementia are often conditions which account for disproportionately higher healthcare service needs and costs as well as worsen health inequalities.^4,26^ Some disease presentations have clear adverse sequelae such as heart failure following acute coronary syndrome. However, multiple events can occur during acute hospitalisation and deteriorations not obviously related to the presenting illness are often not identified or followed up e.g. cardiac dysfunction after non-cardiac surgery.^8^ The impact of new long-term disease and multimorbidity on worsening quality of life and functional status particularly for working age populations is increased for deprived and minority ethnic communities.^27^

### Causal mechanisms

Understanding the causal determinants of these findings is crucial to designing effective public health interventions. Our findings demonstrate imbalances in acute hospitalisation and acquisition of long-term disease by levels of deprivation and ethnic background. Using a holistic approach and addressing psychosocial aspects of health throughout the life course can help to mitigate the wider social determinants of health driving inequalities.^1^ However, disparities in health systems and delivery of services need to be addressed.^4^ Provision of primary care capability continues to be compromised due to workforce gaps in the most deprived areas of the UK.^28^ Ethnic minority patients in England consistently report lower satisfaction with their primary health care as a result of mainly service related factors.^29^ It may be that these factors mean disadvantaged communities are more likely to present to secondary care at a point of acute decline in baseline health rather than seek primary care input with less severe illness. Diagnosis and management of long-term conditions is also more challenging in deprived areas and in people of different minority ethnic backgrounds due to reasons such as increased prevalence of both basic and complex multimorbidity and disparities when relying on self-reporting.^30^ Assessment of socioeconomic status tend to use aggregated measures of area deprivation from multiple components of which, different aspects such as housing will have different levels of relevance across ethnic groups without accurately reflecting cultural norms and life circumstances.^19^ Similarly, ethnicity categorisations are aggregated and do not fully reflect the vast heterogeneity within ethnic categories. Furthermore, multiple interacting dimensions such as gender, social capital, and structural factors including housing, education, employment need to be considered as intersectionality captures ways inequalities result in differential effects based on individual circumstances. To better understand the reasons behind these inequalities, analysis of access to primary care, severity of illness, provision and receipt of both preventative care and treatments, waiting list times for non-acute secondary services, as well as qualitative analysis of experiences and attitudes of the population are required.

### Strengths and limitations

This study has used a large, high quality national dataset including all regions of England and is broadly representative of the general population. We followed a pre-specific statistical analysis plan and included all non-maternity acute hospitalisations over an 11-year period. We were able to evaluate healthcare use across primary and secondary care, consider several baseline sociodemographic and health status risk factors, and examine to an extent, interactions between socioeconomic status and ethnicity. However, there are some limitations. Like many datasets, ethnic categorisations were aggregated and do not reflect the considerable heterogeneity within ethnic groups. A small proportion of patients were classed as unknown or undisclosed ethnicity. Misclassification of patients into both unknown or ‘other’ ethnic groups may occur more frequently in emergency consultations and short-term registrations where follow up data is incomplete leading to the observed markedly lower rates of hospitalisation in these groups. While this data is presented for comparison, strong inferences about these ill-defined ethnic categories should not be drawn as assignment to these groups is likely to be biased toward patients with less clinical contact. Similarly, socioeconomic deprivation has been assessed using a composite measure. Therefore, we could not evaluate the direct effects of variations in other social determinants of health, differences in access to appropriate healthcare, or follow-up services. There may also have been changes in IMD as people age and changes in how people identify by ethnic category over time which were not able to be assessed. We restricted our analysis to acute hospitalisations within the study period and did not analyse ‘first ever hospitalisation’ in life as these cannot be accurately defined and would not reflect contemporary healthcare usage. Reliance on clinical coding meant that assessment of important additional variables including occupation, lifestyle risk factors, variations in disease severity on admission, long-term disease management, and hospital process measures was not possible. Area-level definitions of socioeconomic deprivation may underestimate the effect of inequalities compared to those observed at the individual level and do not account for duration of exposure to specific socioeconomic conditions. We were also not able to include individuals who were not registered with a GP or have a permanent address who are likely to have very high levels of deprivation. Finally, using observational data alone, we provide strong evidence for the association between but are not able prove a causal link between hospitalisation and onset of long-term disease. We did not control for IMD and ethnicity when examining the association between hospitalisation and onset of long-term disease as the causative framework for these relationships is not well established. Inclusion of these variables in regression analyses may attenuate the effects observed related to hospitalisation as a key actionable exposure. Accordingly, we believe our analysis describing differences observed are important and highlights the acute event with healthcare contact as an opportunity for intervention.

### Conclusions

Acute illness and hospitalisation are strongly associated with adverse health outcomes. People in more deprived socioeconomic groups and from minority ethnic backgrounds experience more acute illness despite being a younger age reflecting in a shorter healthy life expectancy. These population inequalities in acute illness are therefore an important driver of inequalities in long-term health. The diversity of acute illness presentations and contributing factors highlights the importance of targeted and sustained public health efforts to reduce inequalities and improve health status for the whole population. Contact with healthcare during acute illness may allow both recognition and management of previously undiagnosed long-term diseases as well as identify opportunities for primary and secondary prevention of new long-term diseases. Acute hospitalisation is likely to represent a crucial but currently underutilised opportunity to change healthy life trajectory and long-term health outcomes. Consideration of healthy life expectancy in the context of secondary care may provide pathways for secondary prevention that have not previously been utilised and additional methods to improve societal health.

## Author statements

### Contributors

All authors were involved in study design, data application, and had access to all data in the study. YIW performed the statistical analysis, which was quality checked by RMP and JRP. All authors contributed to interpretation of the findings. YIW drafted the manuscript. All authors critically revised and approved the final manuscript for submission. The corresponding author attests that all listed authors meet authorship criteria and that no others meeting the criteria have been omitted. YIW is the guarantor. Transparency: The lead author (the guarantor) affirms that the manuscript is an honest, accurate, and transparent account of the study being reported; that no important aspects of the study have been omitted; and that any discrepancies from the study as planned (and, if relevant, registered) have been explained.

### Ethics approval

This study involved secondary use of electronic healthcare records collected as part of routine clinical practice. Access to data was approved via the CPRD research data governance process and this study was conducted in accordance with a pre-specified protocol (CPRD study protocol: 21_000720).

### Provenance and peer review

Not commissioned; externally peer reviewed.

### Data availability statement

All data supporting the findings in this study are provided by CPRD. Access to CPRD data is subject to approval by application via CPRD’s Research Data Governance process.

### Data statements

This research utilised Queen Mary’s Apocrita HPC facility, supported by QMUL Research-IT (doi:10.5281/zenodo.438045).

The CPRD Ethnicity Record sources underlying data from Hospital Episode Statistics (HES) Copyright © 2025, re-used with the permission of The Health & Social Care Information Centre. All rights reserved.

## References

1. Marmot M. Health equity in England: the Marmot review 10 years on. BMJ 2020: m693.

2. Soto GJ, Martin GS, Gong MN. Healthcare disparities in critical illness. Crit Care Med 2013; 41(12): 2784–93.

3. Office of National Statistics (ONS). Deaths involving COVID-19, England and Wales, provisional: week ending 26 March 2021 (Online). 8 April 2021 (Cited: 23 May 2020). Available from: https://www.ons.gov.uk/releases/deathsregisteredweeklyinenglandandwalesprovisionalweekending26march2021 (accessed 12 August 2025).

4. Gkiouleka A, Wong G, Sowden S, et al. Reducing health inequalities through general practice. Lancet Public Health 2023; 8(6): e463–e72.

5. Apea VJ, Wan YI, Dhairyawan R, et al. Ethnicity and outcomes in patients hospitalised with COVID-19 infection in East London: an observational cohort study. BMJ Open 2021; 11(1): e042140.

6. Wan YI, Apea VJ, Dhairyawan R, et al. Ethnic disparities in hospitalisation and hospital-outcomes during the second wave of COVID-19 infection in east London. medRxiv 2021: 2021.07.05.21260026.

7. Wan YI, Robbins AJ, Apea VJ, et al. Ethnicity and acute hospital admissions: Multi-center analysis of routine hospital data. EClinicalMedicine 2021;39: 101077.

8. Ostermann M, Lumlertgul N, Jeong R, See E, Joannidis M, James M. Acute kidney injury. The Lancet 2025; 405(10474): 241–56.

9. The Clinical Practice Research Datalink. The Clinical Practice Research Datalink: primary care data for public health research. 2025. https://cprd.com/primary-care-data-public-health-research (accessed 2 July 2025).

10. NHS Digital. Hospital Episode Statistics. https://scholar.google.com/scholar?q=NHS%20Digital.%20Hospital%20Episode%20statistics.

11. Shiekh SI, Harley M, Ghosh RE, et al. Completeness, agreement, and representativeness of ethnicity recording in the United Kingdom’s Clinical Practice Research Datalink (CPRD) and linked Hospital Episode Statistics (HES). Population Health Metrics 2023; 21(1): 3.

12. Quan H, Sundararajan V, Halfon P, et al. Coding algorithms for defining comorbidities in ICD-9-CM and ICD-10 administrative data. Med Care 2005; 43(11): 1130–9.

13. Kuan V, Denaxas S, Patalay P, et al. Identifying and visualising multimorbidity and comorbidity patterns in patients in the English National Health Service: a population-based study. Lancet Digit Health 2023; 5(1): e16–e27.

14. Hudak D, Johnson D, A. C, et al. Open OnDemand: A web-based client portal for HPC centers. Journal of Open Source Software. The Journal of Open Source Software 2018; 3(27): 622.

15. Lusk JB, Blass B, Mahoney H, et al. Neighborhood socioeconomic deprivation, healthcare access, and 30-day mortality and readmission after sepsis or critical illness: findings from a nationwide study. Crit Care 2023; 27(1): 287.

16. McHenry RD, Moultrie CE, Corfield AR, Lone NI, Mackay DF, Pell JP. The association between socioeconomic status and outcomes in critical illness: A national cohort study of emergency admissions to critical care units in Scotland 2010-2021. J Intensive Care Soc 2025: 17511437251338608.

17. Darlington-Pollock F, Norman P. Examining ethnic inequalities in health and tenure in England: A repeated cross-sectional analysis. Health & Place 2017; 46: 82–90.

18. Allik M, Brown D, Dundas R, Leyland AH. Differences in ill health and in socioeconomic inequalities in health by ethnic groups: a cross-sectional study using 2011 Scottish census. Ethn Health 2019: 1–19.

19. NHS Digita. Hospital Accident & Emergency Activity, 2023-24. https://digital.nhs.uk/data-and-information/publications/statistical/hospital-accident--emergency-activity/2023-24 (accessed 2 July 2025).

20. Public Health Scotland. Acute hospital activity and NHS beds information (annual), Year ending 31 March 2024. https://publichealthscotland.scot/publications/acute-hospital-activity-and-nhs-beds-information-annual/acute-hospital-activity-and-nhs-beds-information-annual-year-ending-31-march-2024/ (accessed 2 July 2025).

21. NHS Digital. Hospital Admitted Patient Care Activity, 2023-24. https://digital.nhs.uk/data-and-information/publications/statistical/hospital-admitted-patient-care-activity/2023-24 (accessed 2 July 2025).

22. Deer RR, Dickinson JM, Baillargeon J, Fisher SR, Raji M, Volpi E. A Phase I Randomized Clinical Trial of Evidence-Based, Pragmatic Interventions to Improve Functional Recovery After Hospitalization in Geriatric Patients. J Gerontol A Biol Sci Med Sci 2019; 74(10): 1628–36.

23. Hashem MD, Nallagangula A, Nalamalapu S, et al. Patient outcomes after critical illness: a systematic review of qualitative studies following hospital discharge. Critical Care 2016; 20(1): 345.

24. Lone NI, Gillies MA, Haddow C, et al. Five-Year Mortality and Hospital Costs Associated with Surviving Intensive Care. Am J Respir Crit Care Med 2016; 194(2): 198–208.

25. Steel N, Ford JA, Newton JN, et al. Changes in health in the countries of the UK and 150 English Local Authority areas 1990&#x2013;2016: a systematic analysis for the Global Burden of Disease Study 2016. The Lancet 2018; 392(10158): 1647–61.

26. Botto F, Alonso-Coello P, Chan MT, et al. Myocardial injury after noncardiac surgery: a large, international, prospective cohort study establishing diagnostic criteria, characteristics, predictors, and 30-day outcomes. Anesthesiology 2014; 120(3): 564–78.

27. Bisquera A, Turner EB, Ledwaba-Chapman L, et al. Inequalities in developing multimorbidity over time: A population-based cohort study from an urban, multi-ethnic borough in the United Kingdom. The Lancet Regional Health – Europe 2022; 12.

28. Armstrong MJ, Wildman JM, Sowden S. How to address the inverse care law and increase GP recruitment in areas of socioeconomic deprivation: a qualitative study of GP trainees’ views and experiences in the UK. BJGP Open 2024; 8(2).

29. Magadi JP, Magadi MA. Ethnic inequalities in patient satisfaction with primary health care in England: Evidence from recent General Practitioner Patient Surveys (GPPS). PLoS One 2022; 17(12): e0270775.

30. Head A, Fleming K, Kypridemos C, Schofield P, Pearson-Stuttard J, O’Flaherty M. Inequalities in incident and prevalent multimorbidity in England, 2004-19: a population-based, descriptive study. The Lancet Healthy Longevity 2021; 2(8): e489-e97.

