## Supplementary file for "Impact of acute hospitalisation on development of long-term disease and health inequality: a longitudinal population study"

Yize I Wan PhD,^1,2*^ Rupert M Pearse MD,^1,2^ John R Prowle MD,^1,2^

^1^William Harvey Research Institute, Queen Mary University of London, London, UK, EC1M 6BQ.

^2^Acute Critical Care Research Unit, Royal London Hospital, Barts Health NHS Trust, London, UK, E1 1FR.

*Correspondence to

Yize I Wan PhD,

Adult Critical Care Unit,

The Royal London Hospital,

Whitechapel

London E1 1FR

**Supplementary file**

Definitions of long-term disease

- Table S1. Definitions of long-term disease.

Cohort diagram

- Figure S1. Cohort definition.

Comparison of CPRD GOLD and Aurum

- Table S2. Comparison of baseline characteristics for patients within CPRD GOLD and Aurum.
- Figure S2. Comparison of IMD and ethnicity distribution for patients within CPRD GOLD and Aurum.
- Figure S3. Comparison of numbers of acute hospitalisations over the study period within CPRD GOLD and Aurum.

Indication for hospitalisation

- Table S3. Main admitting specialty.

Distribution of ethnic groups by IMD

- Figure S4. Distribution of ethnic groups by IMD decile (1 most deprived).

Age-standardised hospitalisation and comparative hospitalisation rates

- Figure S5. Age-standardised hospitalisation rate per 100,000 population per year and comparative hospitalisation ratio by IMD decile, plots by year.
- Table S4. Age-standardised hospitalisation rate per 100,000 population per year and comparative hospitalisation ratio by IMD decile and ethnicity.

Cox proportional hazards modelling

- Table S5. Association of acute hospitalisation with development of long-term disease using Cox proportional hazards modelling.
- Figure S6. Sensitivity analysis time to event curves comparing acute hospitalisation.

Secondary outcomes

- Table S6. Secondary outcomes: total numbers of hospitalisations and multiple hospitalisations by IMD decile and ethnicity.

Healthcare contact days

- Figure S7. Change in healthcare contact days comparing the year before and after acute hospitalisation.
- Figure S8. Change in healthcare contact days comparing the year before and after acute hospitalisation comparing IMD deciles.
- Figure S9. Change in healthcare contact days comparing the year before and after acute hospitalisation comparing ethnic groups.

Age at first hospitalisation

- Table S7. Age at which 50% of patients had experienced hospitalisation by IMD decile and ethnicity.
- Figure S10. Proportion of patients who had hospital admission during the preceding eight years by age in 2021.

RECORD statement

**Table S1. Definition of long-term disease and baseline risk factors.** CHD: coronary heart disease, HF: heart failure, PVD: peripheral vascular disease, PUD: peptic ulcer disease, CVD: cerebrovascular disease.

| **Disease category** | **Included conditions** |
| --- | --- |
| CHD | Coronary heart disease not otherwise specified |
| HF | Heart failure |
| PVD | Peripheral arterial disease |
| CVD | Ischaemic stroke, Stroke not otherwise specified, Transient ischaemic attack |
| Dementia | Dementia |
| Pulmonary | Bronchiectasis, COPD |
| PUD | Gastritis, Duodenitis, Gastro-oesophageal reflux disease, Peptic ulcer |
| Liver | Alcoholic liver disease, Chronic viral hepatitis, Fatty Liver, Hepatic failure |
| Diabetes | Diabetes |
| Renal | Chronic kidney disease, End stage renal disease |
| Cancer | Primary Malignancy: Biliary, Bladder, Bone, Bowel, Brain, Breast, Cervix, Multiple Sites, Oesophageal, Oropharyngeal, Kidney, Liver, Lung, Melanoma, Mesothelioma, Other, Ovary, Pancreas, Prostate, Skin, Stomach, Testis, Thyroid, Uterus, Adrenal. Secondary Malignancy: Bone, Bowel, Brain, Liver, Lung, Lymph Nodes, Peritoneum, Pleura |
| **Risk factor** | **Included conditions** |
| Smoking | Smoking |
| Alcohol | Alcohol misuse |
| Obesity | Obesity |

References for phenotyping algorithms available online:

Kuan V, Denaxas S, Gonzalez-Izquierdo A, et al. A chronological map of 308 physical and mental health conditions from 4 million individuals in the English National Health Service. *The Lancet Digital Health* 2019; **1**(2): e63-e77.

Denaxas SC, George J, Herrett E, et al. Data resource profile: cardiovascular disease research using linked bespoke studies and electronic health records (CALIBER). *Int J Epidemiol* 2012; **41**(6): 1625-38.

Wright AK, Kontopantelis E, Emsley R, et al. Cardiovascular Risk and Risk Factor Management in Type 2 Diabetes Mellitus: A Population-Based Cohort Study Assessing Sex Disparities. *Circulation* 2019; **139**(24): 2742-53.

**Figure S1. Cohort definition.** CPRD: clinical practice research datalink, HES: hospital episode statistics.

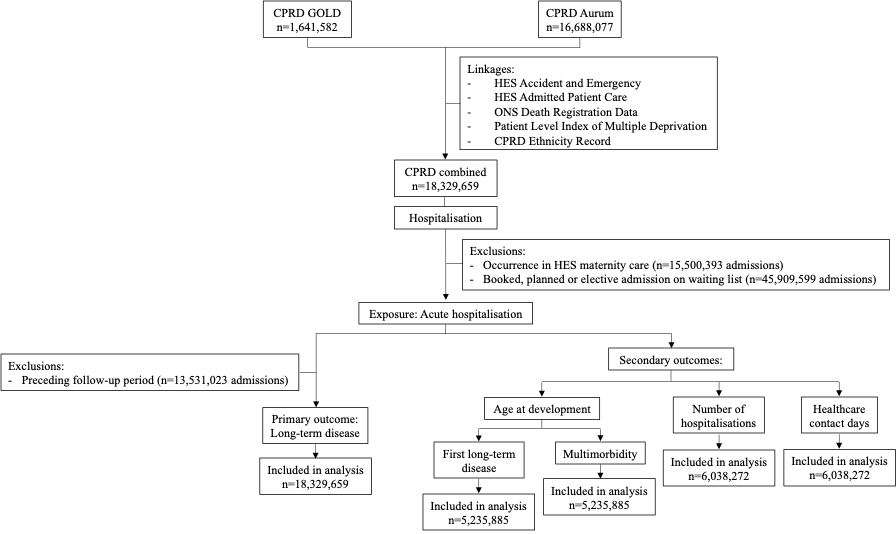

**Table S2. Comparison of baseline characteristics for patients within CPRD GOLD and Aurum.** CHD: coronary heart disease, HF: heart failure, PVD: peripheral vascular disease, PUD: peptic ulcer disease, CVD: cerebrovascular disease. All data presented as n (%) unless otherwise indicated.

|  | **CPRD GOLD** | **CPRD Aurum** |
| --- | --- | --- |
| n | 1641582 | 16688077 |
| *Age in years* |  |  |
| median (IQR) | 43 (28 to 59) | 36 (23 to 53) |
| *Sex (%)* |  |  |
| Male | 804929 (49.0) | 8184947 (49.0) |
| Female | 836634 (51.0) | 8502760 (51.0) |
| Unknown | 19 (0.0) | 370 (0.0) |
| *Smoking (%)* |  |  |
| Current | 42609 (2.6) | 373272 (2.2) |
| Ex | 12331 (0.8) | 100143 (0.6) |
| Never | 12208 (0.7) | 94193 (0.6) |
| Unknown | 30007 (1.8) | 260855 (1.6) |
| *Alcohol intake (%)* | 9506 (0.6) | 74467 (0.4) |
| *Obesity (%)* | 30205 (1.8) | 244919 (1.5) |

**Figure S2. Comparison of IMD and ethnicity distribution for patients within CPRD GOLD and Aurum.** IMD: index of multiple deprivation (1 most deprived).

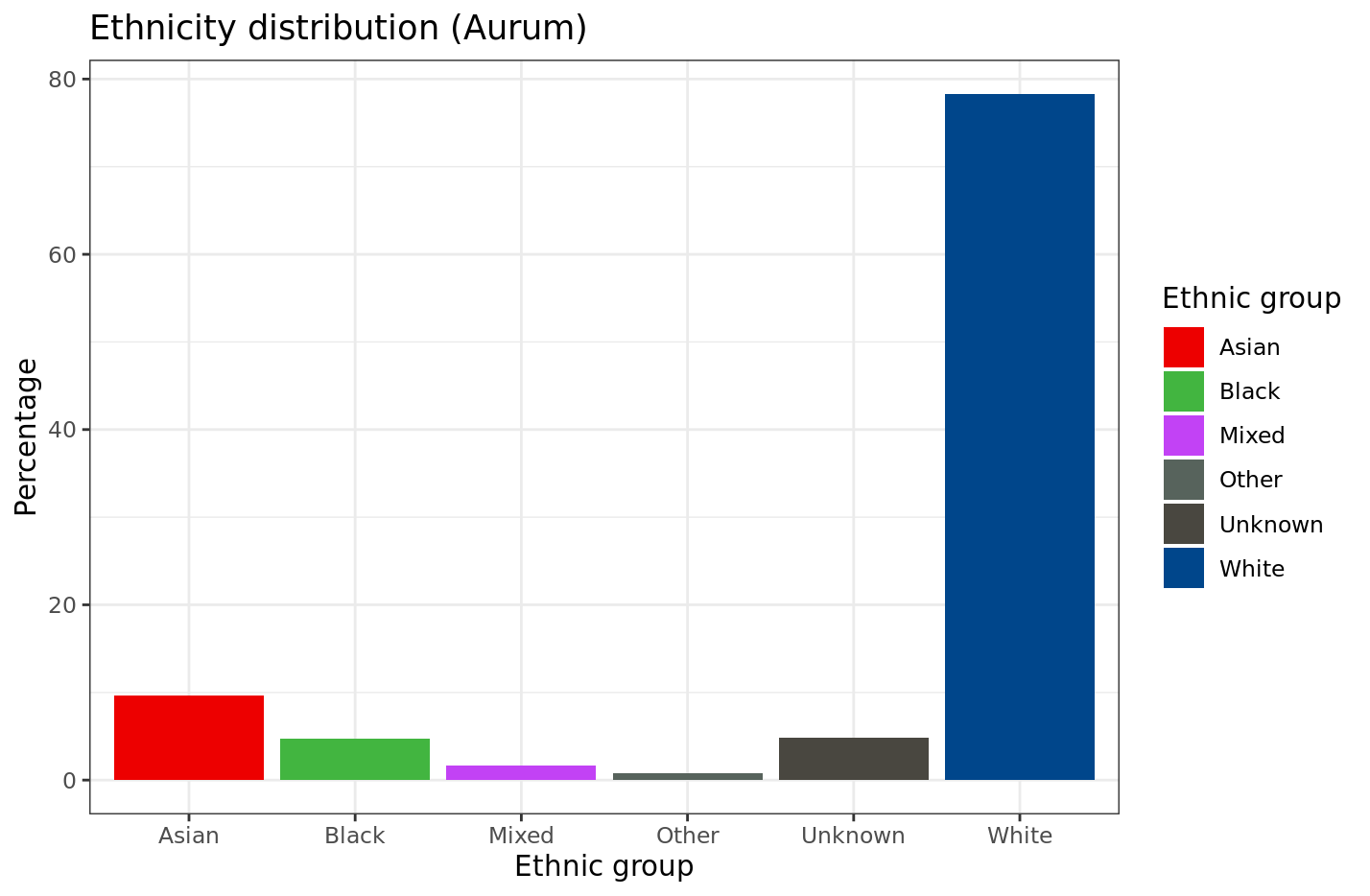

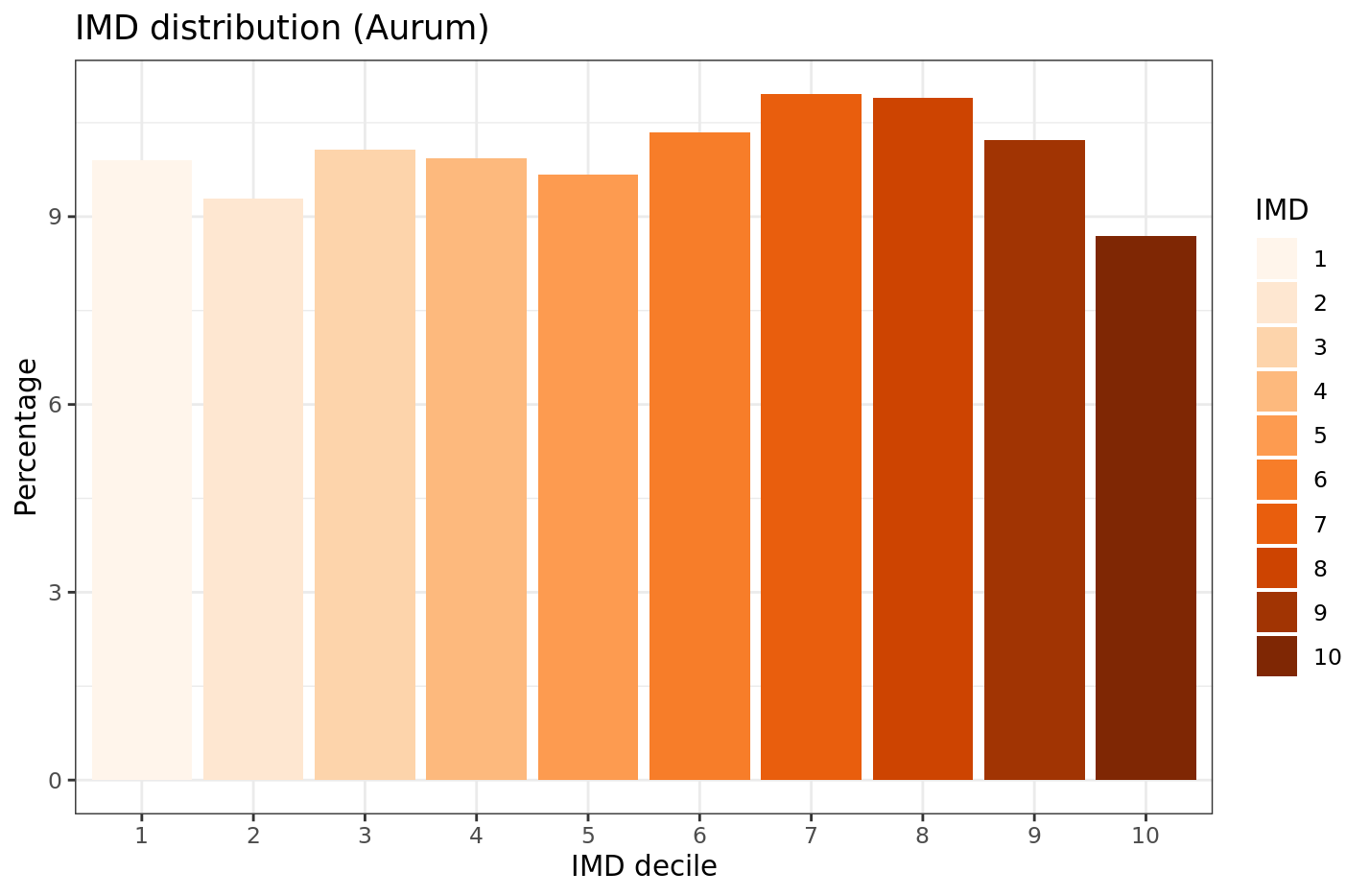

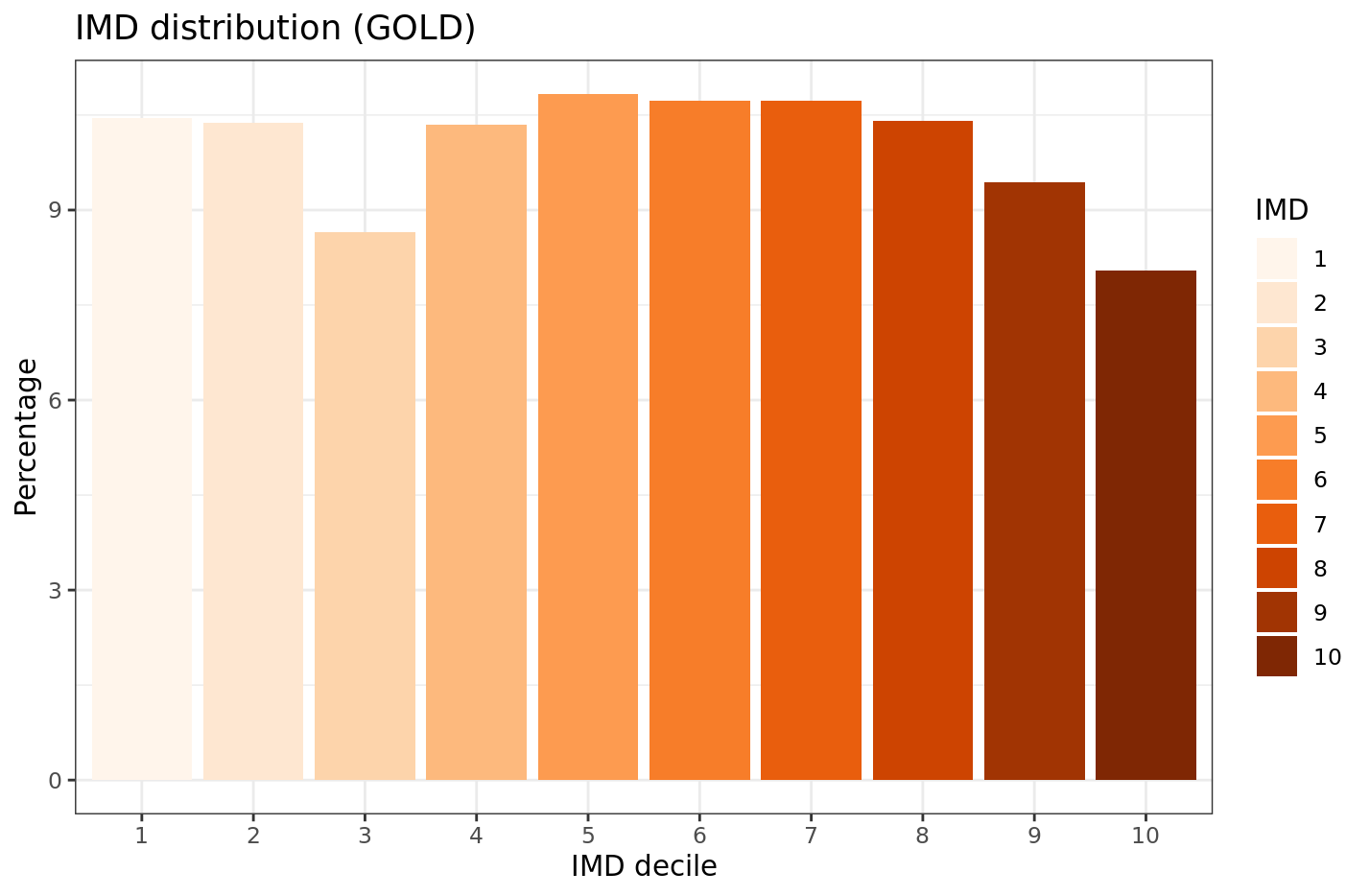

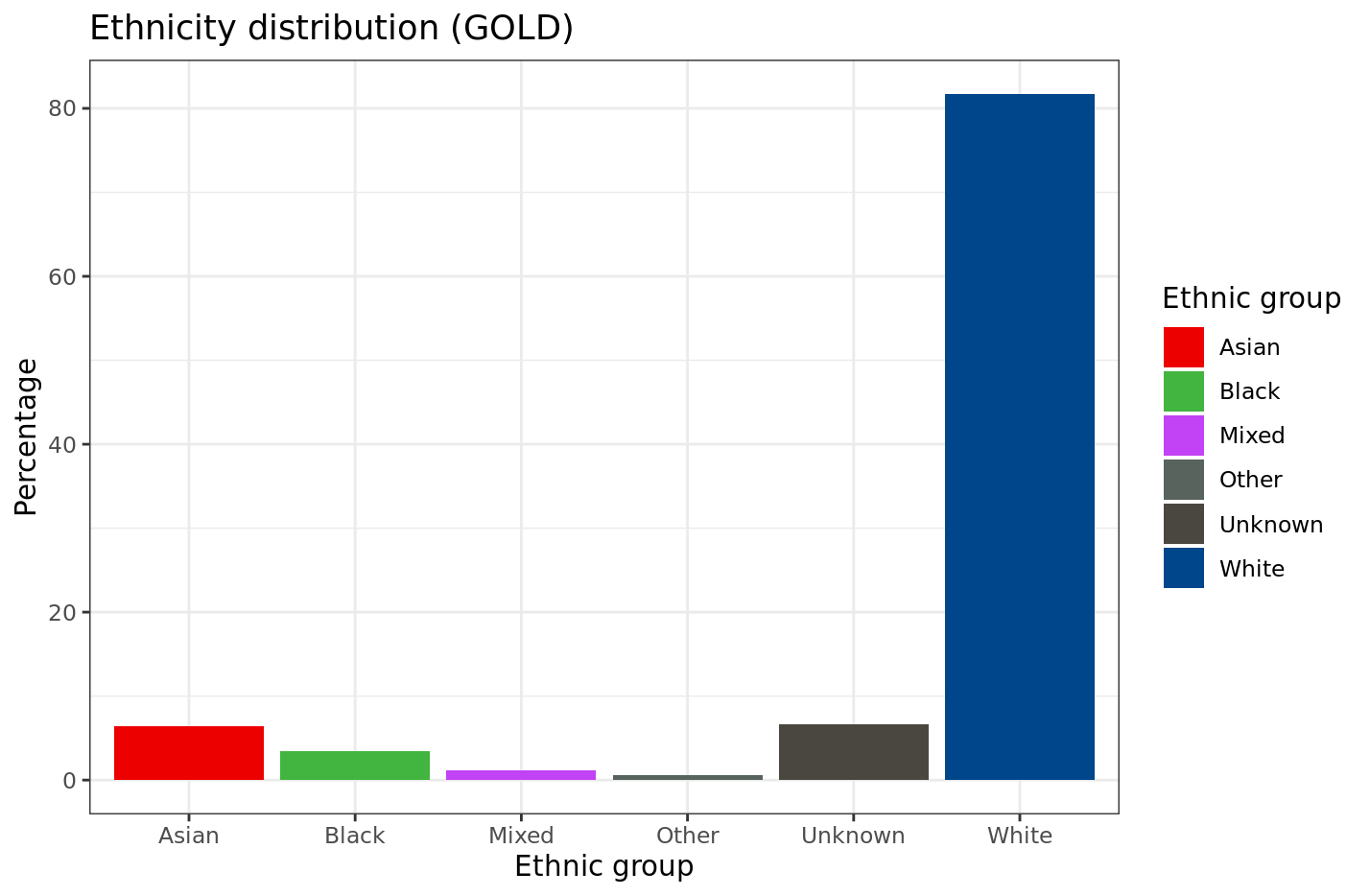

**Figure S3. Comparison of first acute hospitalisations over time within CPRD GOLD and Aurum.** 2021 data to 31st March only.

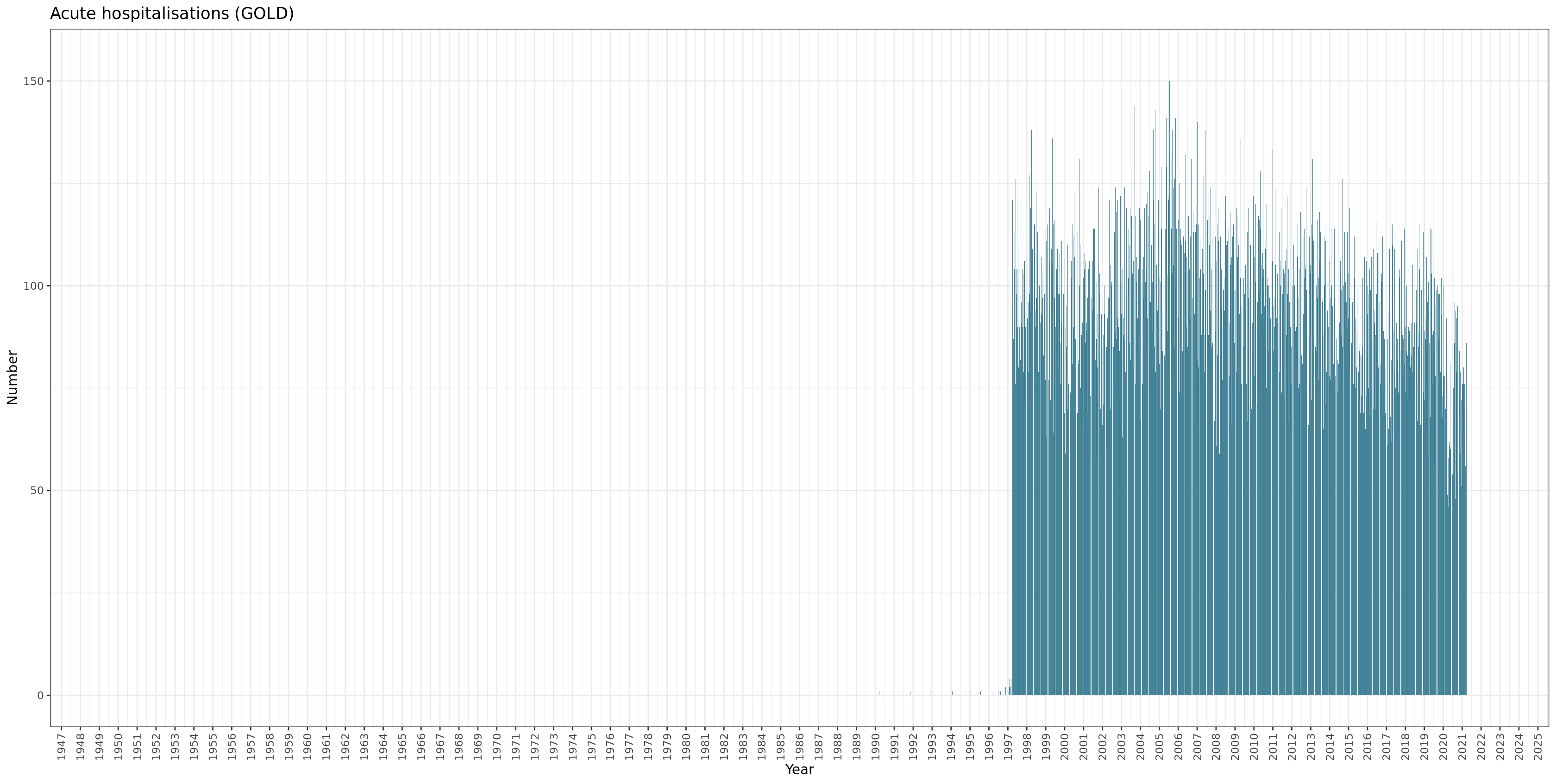

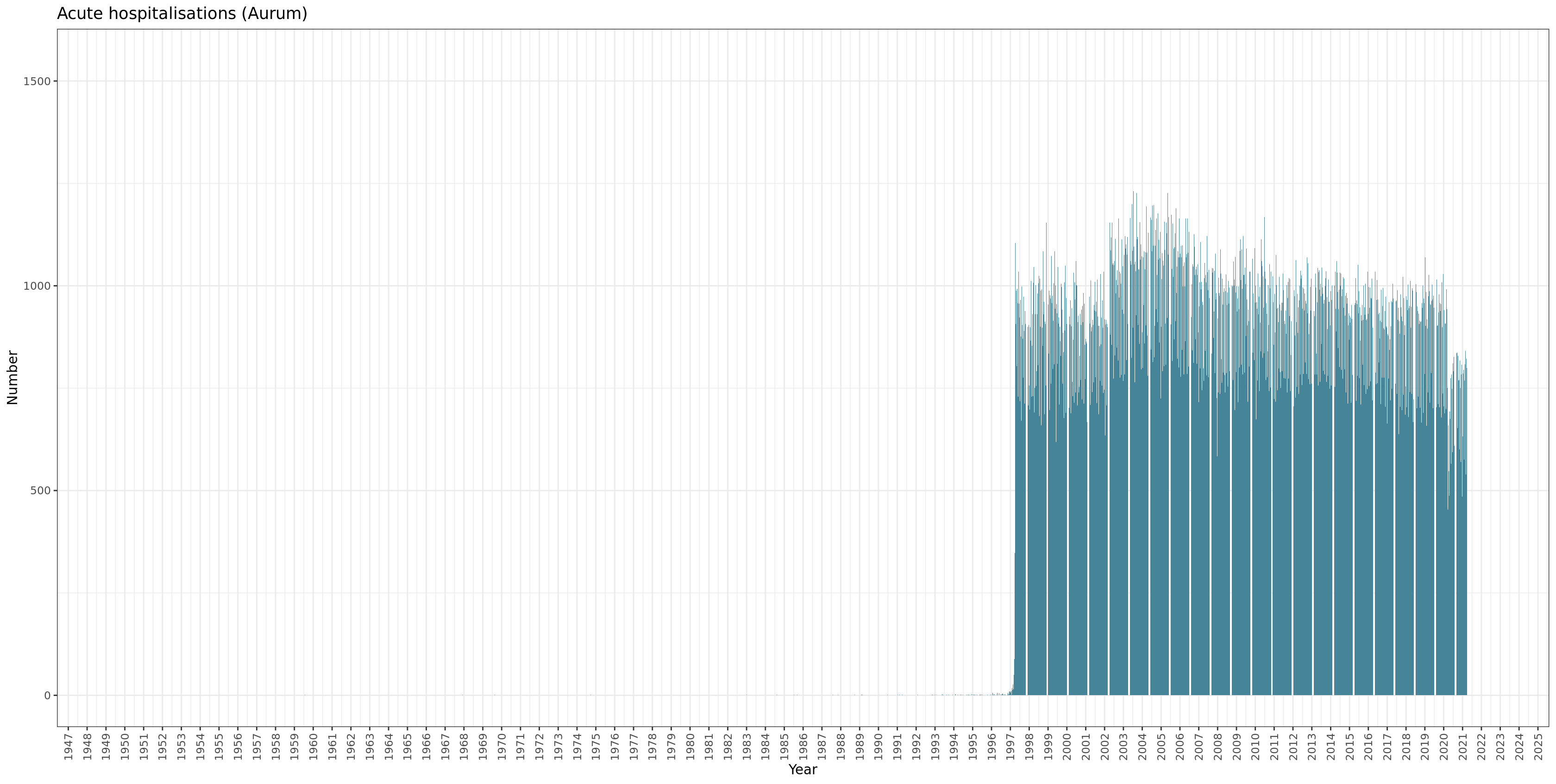

**Table S3. Main admitting specialty.** Note Critical care refers to primary admitting specialty rather than all admissions to the critical care unit including those under another specialty.

| **Specialty** | | **Admissions n (%)** |
| --- | --- | --- |
| Anaesthesia and pain | | 1547 (0.03) |
| Critical care | | 10664 (0.2) |
| Infectious diseases | | 17545 (0.3) |
| Internal medicine | Total | 3405116 (56.4) |
|  | Cardiology | 232059 (3.8) |
|  | Endocrinology | 30040 (0.5) |
|  | Gastroenterology | 93686 (1.6) |
|  | General | 1648017 (27.3) |
|  | Haematology | 19165 (0.3) |
|  | Hepatology | 3366 (0.1) |
|  | Neurology | 28834 (0.5) |
|  | Oncology | 16515 (0.3) |
|  | Other | 1142564 (18.9) |
|  | Renal | 24072 (0.4) |
|  | Respiratory | 136976 (2.3) |
|  | Stroke | 46337 (0.8) |
| Interventional radiology | | 1140 (0.02) |
| Obstetrics and gynaecology | | 875146 (14.5) |
| Oncology | | 15965 (0.3) |
| Psychiatry | | 62849 (1.0) |
| Surgery | Total | 1639505 (27.2) |
|  | Cardiothoracic | 3335 (0.1) |
|  | Colorectal | 50231 (0.8) |
|  | Ear, Nose and Throat | 114381 (1.9) |
|  | General | 704338 (11.7) |
|  | Hepatobiliary and Pancreatic | 5768 (0.1) |
|  | Neurosurgery | 31497 (0.5) |
|  | Ophthalmology | 24917 (0.4) |
|  | Other | 163830 (2.7) |
|  | Transplant | 1687 (0.0) |
|  | Trauma and Orthopaedic | 388429 (6.4) |
|  | Urology | 133386 (2.2) |
|  | Vascular Surgery | 17706 (0.3) |
| Therapies (physiotherapy, speech therapy, dietetics, psychology) | | 3380 (0.06) |
| Unknown | | 5415 (0.09) |

**Figure S4. Distribution of ethnic groups by IMD decile (1 most deprived).**
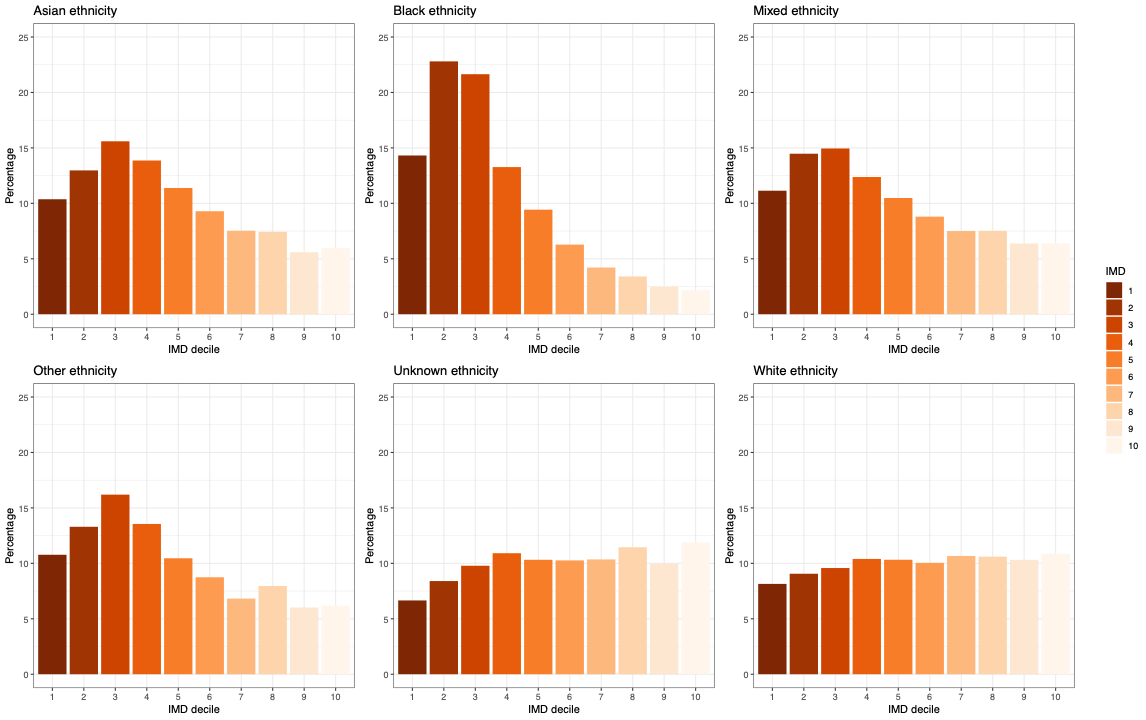

**Figure S5. Age-standardised hospitalisation rate per 100,000 population per year and comparative hospitalisation ratio by IMD decile.** Bar charts showing the age-standardised hospitalisation rate (AHR) per year using ONS population data by Index of Multiple Deprivation decile (IMD 1 most deprived), axis on the left-hand side of each plot. Line graph showing the comparative hospitalisation ratio (CHR) for each IMD decile compared to IMD 10 (least deprived), axis on the right-hand side of each plot. 95% CI shown by error bars. Categories compared in each plot are listed in the legend on the right of each plot. 2021 data to 31st March only.
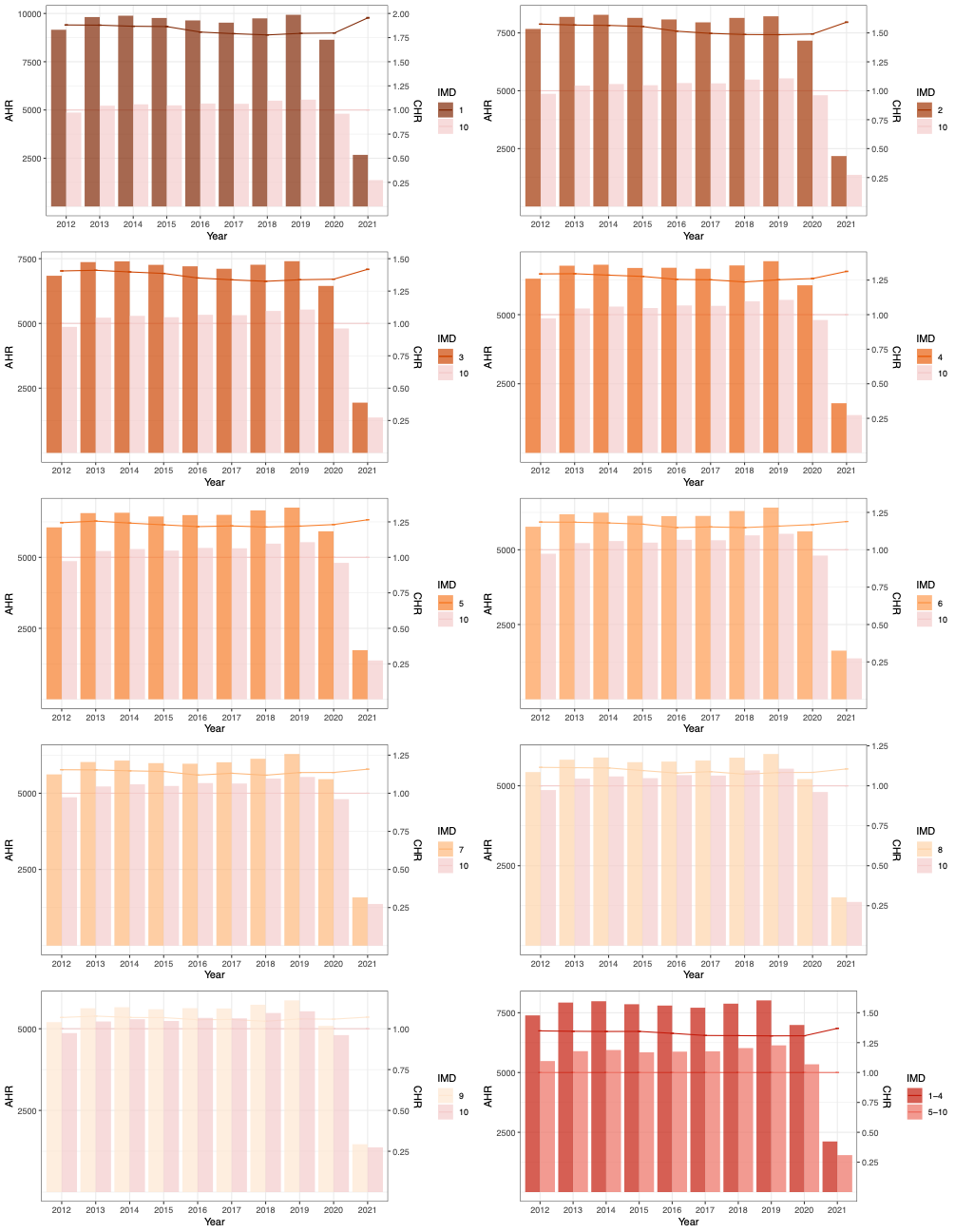

**Table S4. Age-standardised hospitalisation rate per 100,000 population per year and comparative hospitalisation ratio by IMD decile and ethnicity.** AHR: age-standardised hospitalisation rate, CHR: comparative hospitalisation ratio, L95 and U95: lower and upper 95th centile confidence intervals.

| **Year** | **IMD** | **AHR** | **L95 AHR** | **U95 AHR** | **CHR** | **L95 CHR** | **U95 CHR** | **Ethnicity** | **AHR** | **L95 AHR** | **U95 AHR** | **CHR** | **L95 CHR** | **U95 CHR** |
| --- | --- | --- | --- | --- | --- | --- | --- | --- | --- | --- | --- | --- | --- | --- |
| 2012 | 1 | 9156.36 | 9105.22 | 9207.51 | 1.88 | 1.88 | 1.88 | Asian | 6123.88 | 6077.15 | 6170.61 | 0.91 | 0.91 | 0.92 |
|  | 2 | 7671.97 | 7628.00 | 7715.93 | 1.58 | 1.57 | 1.58 | Black | 7205.48 | 7138.77 | 7272.19 | 1.07 | 1.07 | 1.08 |
|  | 3 | 6841.64 | 6801.06 | 6882.23 | 1.41 | 1.40 | 1.41 | Mixed | 5567.02 | 5467.67 | 5666.38 | 0.83 | 0.82 | 0.84 |
|  | 4 | 6302.28 | 6264.10 | 6340.47 | 1.29 | 1.29 | 1.30 | Other | 2005.27 | 1893.39 | 2117.16 | 0.30 | 0.28 | 0.32 |
|  | 5 | 6056.43 | 6018.79 | 6094.07 | 1.24 | 1.24 | 1.25 | Unknown | 1116.04 | 1088.17 | 1143.91 | 0.17 | 0.16 | 0.17 |
|  | 6 | 5771.23 | 5733.56 | 5808.91 | 1.19 | 1.19 | 1.19 | White | 6703.68 | 6689.51 | 6717.86 | 1.00 | 1.00 | 1.00 |
|  | 7 | 5619.19 | 5583.03 | 5655.35 | 1.15 | 1.15 | 1.15 |  | | | | | | |
|  | 8 | 5429.35 | 5393.54 | 5465.16 | 1.12 | 1.12 | 1.12 |  |  |  |  |  |  |  |
|  | 9 | 5204.32 | 5168.84 | 5239.81 | 1.07 | 1.07 | 1.07 |  |  |  |  |  |  |  |
|  | 10 | 4867.74 | 4834.24 | 4901.24 | 1.00 | 1.00 | 1.00 |  |  |  |  |  |  |  |
|  | 1-4 | 7389.14 | 7367.64 | 7410.63 | 1.35 | 1.35 | 1.35 |  |  |  |  |  |  |  |
|  | 5-10 | 5480.11 | 5465.42 | 5494.80 | 1.00 | 1.00 | 1.00 |  |  |  |  |  |  |  |
| 2013 | 1 | 9814.85 | 9762.01 | 9867.69 | 1.88 | 1.88 | 1.88 | Asian | 6588.75 | 6539.66 | 6637.84 | 0.91 | 0.91 | 0.92 |
|  | 2 | 8191.13 | 8145.79 | 8236.47 | 1.57 | 1.57 | 1.57 | Black | 7558.42 | 7489.56 | 7627.27 | 1.05 | 1.04 | 1.06 |
|  | 3 | 7368.33 | 7326.36 | 7410.31 | 1.41 | 1.41 | 1.41 | Mixed | 6019.62 | 5915.10 | 6124.14 | 0.83 | 0.82 | 0.85 |
|  | 4 | 6771.19 | 6731.85 | 6810.52 | 1.30 | 1.30 | 1.30 | Other | 2397.10 | 2275.59 | 2518.62 | 0.33 | 0.32 | 0.35 |
|  | 5 | 6559.07 | 6520.18 | 6597.96 | 1.26 | 1.25 | 1.26 | Unknown | 1352.70 | 1321.57 | 1383.83 | 0.19 | 0.18 | 0.19 |
|  | 6 | 6186.68 | 6148.09 | 6225.26 | 1.18 | 1.18 | 1.18 | White | 7214.56 | 7200.00 | 7229.13 | 1.00 | 1.00 | 1.00 |
|  | 7 | 6024.87 | 5987.86 | 6061.87 | 1.15 | 1.15 | 1.15 |  | | | | | | |
|  | 8 | 5816.26 | 5779.65 | 5852.87 | 1.11 | 1.11 | 1.11 |  |  |  |  |  |  |  |
|  | 9 | 5628.91 | 5592.50 | 5665.32 | 1.08 | 1.08 | 1.08 |  |  |  |  |  |  |  |
|  | 10 | 5223.60 | 5189.37 | 5257.82 | 1.00 | 1.00 | 1.00 |  |  |  |  |  |  |  |
|  | 1-4 | 7925.90 | 7903.71 | 7948.09 | 1.34 | 1.35 | 1.34 |  |  |  |  |  |  |  |
|  | 5-10 | 5893.14 | 5878.08 | 5908.20 | 1.00 | 1.00 | 1.00 |  |  |  |  |  |  |  |
| 2014 | 1 | 9884.65 | 9832.42 | 9936.88 | 1.87 | 1.87 | 1.87 | Asian | 6698.88 | 6650.51 | 6747.25 | 0.92 | 0.91 | 0.92 |
|  | 2 | 8282.49 | 8237.66 | 8327.32 | 1.56 | 1.56 | 1.57 | Black | 7598.47 | 7530.97 | 7665.98 | 1.04 | 1.03 | 1.05 |
|  | 3 | 7398.32 | 7357.07 | 7439.57 | 1.40 | 1.40 | 1.40 | Mixed | 6232.54 | 6127.91 | 6337.16 | 0.85 | 0.84 | 0.87 |
|  | 4 | 6806.24 | 6767.55 | 6844.92 | 1.29 | 1.28 | 1.29 | Other | 2222.54 | 2113.71 | 2331.38 | 0.30 | 0.29 | 0.32 |
|  | 5 | 6571.98 | 6533.74 | 6610.22 | 1.24 | 1.24 | 1.24 | Unknown | 1383.55 | 1353.01 | 1414.10 | 0.19 | 0.19 | 0.19 |
|  | 6 | 6242.67 | 6204.58 | 6280.77 | 1.18 | 1.18 | 1.18 | White | 7306.22 | 7291.75 | 7320.68 | 1.00 | 1.00 | 1.00 |
|  | 7 | 6071.98 | 6035.44 | 6108.51 | 1.15 | 1.15 | 1.15 |  | | | | | | |
|  | 8 | 5884.00 | 5847.81 | 5920.20 | 1.11 | 1.11 | 1.11 |  |  |  |  |  |  |  |
|  | 9 | 5659.91 | 5624.01 | 5695.82 | 1.07 | 1.07 | 1.07 |  |  |  |  |  |  |  |
|  | 10 | 5293.17 | 5259.29 | 5327.04 | 1.00 | 1.00 | 1.00 |  |  |  |  |  |  |  |
|  | 1-4 | 7981.47 | 7959.59 | 8003.34 | 1.34 | 1.34 | 1.34 |  |  |  |  |  |  |  |
|  | 5-10 | 5941.60 | 5926.73 | 5956.46 | 1.00 | 1.00 | 1.00 |  |  |  |  |  |  |  |
| 2015 | 1 | 9767.20 | 9715.88 | 9818.52 | 1.86 | 1.86 | 1.87 | Asian | 6598.86 | 6551.65 | 6646.07 | 0.91 | 0.91 | 0.92 |
|  | 2 | 8149.93 | 8106.03 | 8193.83 | 1.56 | 1.55 | 1.56 | Black | 7564.24 | 7497.63 | 7630.84 | 1.05 | 1.04 | 1.06 |
|  | 3 | 7262.52 | 7222.20 | 7302.84 | 1.39 | 1.39 | 1.39 | Mixed | 6272.46 | 6168.47 | 6376.45 | 0.87 | 0.86 | 0.88 |
|  | 4 | 6688.86 | 6651.04 | 6726.68 | 1.28 | 1.28 | 1.28 | Other | 2228.38 | 2122.57 | 2334.18 | 0.31 | 0.29 | 0.32 |
|  | 5 | 6440.05 | 6402.73 | 6477.38 | 1.23 | 1.23 | 1.23 | Unknown | 1377.88 | 1348.08 | 1407.69 | 0.19 | 0.19 | 0.19 |
|  | 6 | 6135.45 | 6098.20 | 6172.69 | 1.17 | 1.17 | 1.17 | White | 7214.32 | 7200.10 | 7228.53 | 1.00 | 1.00 | 1.00 |
|  | 7 | 5988.32 | 5952.53 | 6024.12 | 1.14 | 1.14 | 1.14 |  | | | | | | |
|  | 8 | 5739.45 | 5704.22 | 5774.69 | 1.10 | 1.10 | 1.10 |  |  |  |  |  |  |  |
|  | 9 | 5596.79 | 5561.54 | 5632.04 | 1.07 | 1.07 | 1.07 |  |  |  |  |  |  |  |
|  | 10 | 5239.15 | 5205.93 | 5272.37 | 1.00 | 1.00 | 1.00 |  |  |  |  |  |  |  |
|  | 1-4 | 7855.81 | 7834.38 | 7877.24 | 1.34 | 1.34 | 1.34 |  |  |  |  |  |  |  |
|  | 5-10 | 5845.20 | 5830.66 | 5859.74 | 1.00 | 1.00 | 1.00 |  |  |  |  |  |  |  |
| 2016 | 1 | 9640.07 | 9589.60 | 9690.55 | 1.81 | 1.81 | 1.81 | Asian | 6700.18 | 6653.46 | 6746.91 | 0.93 | 0.92 | 0.93 |
|  | 2 | 8081.79 | 8038.65 | 8124.93 | 1.52 | 1.51 | 1.52 | Black | 7468.39 | 7403.25 | 7533.53 | 1.03 | 1.03 | 1.04 |
|  | 3 | 7206.18 | 7166.59 | 7245.77 | 1.35 | 1.35 | 1.35 | Mixed | 6299.42 | 6197.09 | 6401.76 | 0.87 | 0.86 | 0.88 |
|  | 4 | 6695.98 | 6658.61 | 6733.35 | 1.26 | 1.25 | 1.26 | Other | 2343.66 | 2239.10 | 2448.22 | 0.32 | 0.31 | 0.34 |
|  | 5 | 6489.22 | 6452.25 | 6526.19 | 1.22 | 1.22 | 1.22 | Unknown | 1406.78 | 1377.47 | 1436.09 | 0.19 | 0.19 | 0.20 |
|  | 6 | 6126.07 | 6089.33 | 6162.81 | 1.15 | 1.15 | 1.15 | White | 7236.56 | 7222.46 | 7250.66 | 1.00 | 1.00 | 1.00 |
|  | 7 | 5969.49 | 5934.22 | 6004.76 | 1.12 | 1.12 | 1.12 |  | | | | | | |
|  | 8 | 5758.45 | 5723.60 | 5793.30 | 1.08 | 1.08 | 1.08 |  |  |  |  |  |  |  |
|  | 9 | 5629.97 | 5595.06 | 5664.89 | 1.06 | 1.06 | 1.06 |  |  |  |  |  |  |  |
|  | 10 | 5333.83 | 5300.72 | 5366.93 | 1.00 | 1.00 | 1.00 |  |  |  |  |  |  |  |
|  | 1-4 | 7800.14 | 7779.05 | 7821.23 | 1.33 | 1.33 | 1.33 |  |  |  |  |  |  |  |
|  | 5-10 | 5874.25 | 5859.85 | 5888.65 | 1.00 | 1.00 | 1.00 |  |  |  |  |  |  |  |
| 2017 | 1 | 9524.28 | 9474.62 | 9573.94 | 1.79 | 1.79 | 1.79 | Asian | 6697.37 | 6651.30 | 6743.43 | 0.92 | 0.92 | 0.93 |
|  | 2 | 7956.22 | 7913.89 | 7998.55 | 1.50 | 1.49 | 1.50 | Black | 7470.83 | 7406.22 | 7535.44 | 1.03 | 1.02 | 1.04 |
|  | 3 | 7111.24 | 7072.38 | 7150.10 | 1.34 | 1.34 | 1.34 | Mixed | 6357.57 | 6256.61 | 6458.53 | 0.88 | 0.86 | 0.89 |
|  | 4 | 6660.56 | 6623.73 | 6697.39 | 1.25 | 1.25 | 1.25 | Other | 2630.56 | 2520.66 | 2740.46 | 0.36 | 0.35 | 0.38 |
|  | 5 | 6497.44 | 6460.82 | 6534.06 | 1.22 | 1.22 | 1.22 | Unknown | 1446.11 | 1416.77 | 1475.44 | 0.20 | 0.20 | 0.20 |
|  | 6 | 6131.90 | 6095.55 | 6168.25 | 1.15 | 1.15 | 1.15 | White | 7253.87 | 7239.85 | 7267.89 | 1.00 | 1.00 | 1.00 |
|  | 7 | 6016.80 | 5981.75 | 6051.85 | 1.13 | 1.13 | 1.13 |  | | | | | | |
|  | 8 | 5790.30 | 5755.76 | 5824.83 | 1.09 | 1.09 | 1.09 |  |  |  |  |  |  |  |
|  | 9 | 5621.96 | 5587.42 | 5656.50 | 1.06 | 1.06 | 1.06 |  |  |  |  |  |  |  |
|  | 10 | 5319.20 | 5286.56 | 5351.85 | 1.00 | 1.00 | 1.00 |  |  |  |  |  |  |  |
|  | 1-4 | 7710.22 | 7689.49 | 7730.95 | 1.31 | 1.31 | 1.31 |  |  |  |  |  |  |  |
|  | 5-10 | 5886.55 | 5872.29 | 5900.80 | 1.00 | 1.00 | 1.00 |  |  |  |  |  |  |  |
| 2018 | 1 | 9746.82 | 9696.94 | 9796.69 | 1.78 | 1.78 | 1.78 | Asian | 6996.24 | 6949.83 | 7042.66 | 0.94 | 0.94 | 0.95 |
|  | 2 | 8148.41 | 8105.94 | 8190.89 | 1.49 | 1.49 | 1.49 | Black | 7834.79 | 7769.21 | 7900.36 | 1.05 | 1.05 | 1.06 |
|  | 3 | 7266.39 | 7227.51 | 7305.26 | 1.33 | 1.32 | 1.33 | Mixed | 6830.23 | 6725.54 | 6934.92 | 0.92 | 0.91 | 0.93 |
|  | 4 | 6780.51 | 6743.71 | 6817.30 | 1.24 | 1.24 | 1.24 | Other | 2734.96 | 2630.17 | 2839.75 | 0.37 | 0.35 | 0.38 |
|  | 5 | 6654.90 | 6618.16 | 6691.64 | 1.21 | 1.21 | 1.21 | Unknown | 1536.59 | 1507.04 | 1566.15 | 0.21 | 0.20 | 0.21 |
|  | 6 | 6296.73 | 6260.22 | 6333.23 | 1.15 | 1.15 | 1.15 | White | 7431.01 | 7416.89 | 7445.13 | 1.00 | 1.00 | 1.00 |
|  | 7 | 6132.36 | 6097.26 | 6167.45 | 1.12 | 1.12 | 1.12 |  | | | | | | |
|  | 8 | 5880.85 | 5846.36 | 5915.35 | 1.07 | 1.07 | 1.07 |  |  |  |  |  |  |  |
|  | 9 | 5736.03 | 5701.42 | 5770.63 | 1.05 | 1.05 | 1.05 |  |  |  |  |  |  |  |
|  | 10 | 5482.83 | 5449.95 | 5515.70 | 1.00 | 1.00 | 1.00 |  |  |  |  |  |  |  |
|  | 1-4 | 7879.63 | 7858.86 | 7900.40 | 1.31 | 1.31 | 1.31 |  |  |  |  |  |  |  |
|  | 5-10 | 6021.89 | 6007.60 | 6036.19 | 1.00 | 1.00 | 1.00 |  |  |  |  |  |  |  |
| 2019 | 1 | 9931.29 | 9881.23 | 9981.36 | 1.79 | 1.79 | 1.80 | Asian | 7232.50 | 7186.07 | 7278.93 | 0.95 | 0.95 | 0.96 |
|  | 2 | 8217.62 | 8175.32 | 8259.93 | 1.48 | 1.48 | 1.49 | Black | 8059.12 | 7993.34 | 8124.90 | 1.06 | 1.06 | 1.07 |
|  | 3 | 7403.45 | 7364.51 | 7442.39 | 1.34 | 1.34 | 1.34 | Mixed | 7004.12 | 6899.82 | 7108.41 | 0.92 | 0.91 | 0.94 |
|  | 4 | 6933.85 | 6896.93 | 6970.77 | 1.25 | 1.25 | 1.25 | Other | 3022.60 | 2915.38 | 3129.82 | 0.40 | 0.39 | 0.41 |
|  | 5 | 6752.98 | 6716.22 | 6789.74 | 1.22 | 1.22 | 1.22 | Unknown | 1641.75 | 1611.69 | 1671.80 | 0.22 | 0.21 | 0.22 |
|  | 6 | 6410.66 | 6374.05 | 6447.27 | 1.16 | 1.16 | 1.16 | White | 7578.40 | 7564.19 | 7592.61 | 1.00 | 1.00 | 1.00 |
|  | 7 | 6287.88 | 6252.53 | 6323.24 | 1.14 | 1.14 | 1.14 |  | | | | | | |
|  | 8 | 5994.63 | 5960.03 | 6029.23 | 1.08 | 1.08 | 1.08 |  |  |  |  |  |  |  |
|  | 9 | 5868.86 | 5834.06 | 5903.66 | 1.06 | 1.06 | 1.06 |  |  |  |  |  |  |  |
|  | 10 | 5535.40 | 5502.64 | 5568.16 | 1.00 | 1.00 | 1.00 |  |  |  |  |  |  |  |
|  | 1-4 | 8016.37 | 7995.57 | 8037.18 | 1.31 | 1.31 | 1.31 |  |  |  |  |  |  |  |
|  | 5-10 | 6135.82 | 6121.48 | 6150.16 | 1.00 | 1.00 | 1.00 |  |  |  |  |  |  |  |
| 2020 | 1 | 8639.29 | 8592.79 | 8685.79 | 1.80 | 1.80 | 1.80 | Asian | 6061.93 | 6020.04 | 6103.83 | 0.91 | 0.91 | 0.92 |
|  | 2 | 7165.35 | 7126.01 | 7204.68 | 1.49 | 1.49 | 1.49 | Black | 6869.85 | 6809.67 | 6930.02 | 1.03 | 1.03 | 1.04 |
|  | 3 | 6448.34 | 6412.17 | 6484.52 | 1.34 | 1.34 | 1.34 | Mixed | 6231.20 | 6132.76 | 6329.65 | 0.94 | 0.92 | 0.95 |
|  | 4 | 6061.94 | 6027.60 | 6096.28 | 1.26 | 1.26 | 1.26 | Other | 2991.65 | 2884.56 | 3098.74 | 0.45 | 0.43 | 0.47 |
|  | 5 | 5917.14 | 5882.87 | 5951.40 | 1.23 | 1.23 | 1.23 | Unknown | 1536.03 | 1507.15 | 1564.91 | 0.23 | 0.23 | 0.24 |
|  | 6 | 5615.25 | 5581.14 | 5649.36 | 1.17 | 1.17 | 1.17 | White | 6644.69 | 6631.41 | 6657.96 | 1.00 | 1.00 | 1.00 |
|  | 7 | 5462.21 | 5429.43 | 5494.98 | 1.14 | 1.14 | 1.14 |  | | | | | | |
|  | 8 | 5209.77 | 5177.70 | 5241.84 | 1.08 | 1.08 | 1.08 |  |  |  |  |  |  |  |
|  | 9 | 5089.74 | 5057.52 | 5121.96 | 1.06 | 1.06 | 1.06 |  |  |  |  |  |  |  |
|  | 10 | 4806.49 | 4776.20 | 4836.78 | 1.00 | 1.00 | 1.00 |  |  |  |  |  |  |  |
|  | 1-4 | 6988.39 | 6969.05 | 7007.72 | 1.31 | 1.31 | 1.31 |  |  |  |  |  |  |  |
|  | 5-10 | 5345.15 | 5331.84 | 5358.46 | 1.00 | 1.00 | 1.00 |  |  |  |  |  |  |  |
| 2021 | 1 | 2674.91 | 2649.13 | 2700.68 | 1.96 | 1.95 | 1.96 | Asian | 1904.49 | 1880.94 | 1928.05 | 0.97 | 0.96 | 0.98 |
|  | 2 | 2179.14 | 2157.55 | 2200.73 | 1.59 | 1.59 | 1.60 | Black | 2113.07 | 2079.62 | 2146.52 | 1.08 | 1.06 | 1.09 |
|  | 3 | 1940.16 | 1920.42 | 1959.91 | 1.42 | 1.42 | 1.42 | Mixed | 1894.29 | 1839.66 | 1948.92 | 0.97 | 0.94 | 0.99 |
|  | 4 | 1796.40 | 1777.84 | 1814.97 | 1.31 | 1.31 | 1.31 | Other | 898.19 | 837.70 | 958.67 | 0.46 | 0.43 | 0.49 |
|  | 5 | 1730.49 | 1712.10 | 1748.88 | 1.26 | 1.26 | 1.27 | Unknown | 447.05 | 431.64 | 462.47 | 0.23 | 0.22 | 0.24 |
|  | 6 | 1626.57 | 1608.40 | 1644.74 | 1.19 | 1.19 | 1.19 | White | 1960.31 | 1953.14 | 1967.48 | 1.00 | 1.00 | 1.00 |
|  | 7 | 1584.11 | 1566.62 | 1601.60 | 1.16 | 1.16 | 1.16 |  | | | | | | |
|  | 8 | 1512.60 | 1495.46 | 1529.74 | 1.11 | 1.11 | 1.11 |  |  |  |  |  |  |  |
|  | 9 | 1465.78 | 1448.69 | 1482.87 | 1.07 | 1.07 | 1.07 |  |  |  |  |  |  |  |
|  | 10 | 1368.10 | 1352.14 | 1384.06 | 1.00 | 1.00 | 1.00 |  |  |  |  |  |  |  |
|  | 1-4 | 2117.07 | 2106.48 | 2127.66 | 1.37 | 1.37 | 1.37 |  |  |  |  |  |  |  |
|  | 5-10 | 1546.08 | 1538.99 | 1553.17 | 1.00 | 1.00 | 1.00 |  |  |  |  |  |  |  |

Example calculation for IMD 1 compared to IMD 10 in 2012. AHR: age-standardised hospitalisation rate, CHR: comparative hospitalisation ratio, L95 and U95: lower and upper 95th centile confidence intervals.

| **Year** | **IMD** | **Age group** | **Total** | **Hospitalised** | **Hospitalisation rate** | **Standard population** | **Expected hospitalisation** | **AHR** | **L95 AHR** | **U95 AHR** | **CHR** | **L95 CHR** | **U95 CHR** |
| --- | --- | --- | --- | --- | --- | --- | --- | --- | --- | --- | --- | --- | --- |
| 2012 | 1 | 10-19 | 281562 | 11910 | 4229.97422 | 1355458 | 5733552390 |  |  |  |  |  |  |
|  |  | 20-29 | 368993 | 27516 | 7457.05203 | 7264679 | 54173089290 |  |  |  |  |  |  |
|  |  | 30-39 | 274049 | 18967 | 6921.02507 | 7057901 | 48847909778 |  |  |  |  |  |  |
|  |  | 40-49 | 237942 | 16191 | 6804.59944 | 7785645 | 52978195609 |  |  |  |  |  |  |
|  |  | 50-59 | 173595 | 13232 | 7622.33935 | 6580951 | 50162241788 |  |  |  |  |  |  |
|  |  | 60-69 | 117658 | 11344 | 9641.50334 | 5801133 | 55931643196 |  |  |  |  |  |  |
|  |  | 70-79 | 76582 | 11614 | 15165.4436 | 3753037 | 56916470865 |  |  |  |  |  |  |
|  |  | 80-89 | 42414 | 9745 | 22975.9042 | 2066551 | 47480877764 |  |  |  |  |  |  |
|  |  | 90+ | 8595 | 2610 | 30366.4921 | 437487 | 13284945550 |  |  |  |  |  |  |
|  |  | Total | 1581390 | 123129 | 7786.124865 | 42102842 | 385508926230 | 9156.36351 | 9105.21901 | 9207.50801 | 1.88103063 | 1.88348591 | 1.87860891 |
|  | 10 | 10-19 | 285401 | 6296 | 2206.0189 | 1355458 | 2990165966 |  |  |  |  |  |  |
|  |  | 20-29 | 285419 | 10022 | 3511.32896 | 7264679 | 25508677747 |  |  |  |  |  |  |
|  |  | 30-39 | 287587 | 12647 | 4397.62576 | 7057901 | 31038007263 |  |  |  |  |  |  |
|  |  | 40-49 | 299565 | 8340 | 2784.03685 | 7785645 | 21675522608 |  |  |  |  |  |  |
|  |  | 50-59 | 239574 | 7498 | 3129.72192 | 6580951 | 20596546619 |  |  |  |  |  |  |
|  |  | 60-69 | 205335 | 9258 | 4508.72964 | 5801133 | 26155740285 |  |  |  |  |  |  |
|  |  | 70-79 | 129987 | 10939 | 8415.45693 | 3753037 | 31583521231 |  |  |  |  |  |  |
|  |  | 80-89 | 73313 | 12148 | 16570.049 | 2066551 | 34242851265 |  |  |  |  |  |  |
|  |  | 90+ | 15547 | 3964 | 25496.8804 | 437487 | 11154553727 |  |  |  |  |  |  |
|  |  | Total | 1821728 | 81112 | 4452.475891 | 42102842 | 204945586711 | 4867.73759 | 4834.23791 | 4901.23727 | 1 | 1 | 1 |

**Table S5.** **Association of acute hospitalisation with development of long-term disease using Cox proportional hazards modelling.** *Variables included age, sex (excluding unknown), and baseline number of chronic diseases.

|  | **Model** | **Hazards ratio (95% CI)** | **p value** |
| --- | --- | --- | --- |
| **Main** | Unadjusted  (n=24359616, events=3261464) | 3.52 (3.51 to 3.52) | <0.0001 |
|  | Adjusted*  (n=24359161, events=3261418) | 2.48 (2.47 to 2.48) | <0.0001 |
| **Sensitivity** | Unadjusted  (n=24359616, events=3659346) | 3.78 (3.77 to 3.78) | <0.0001 |
|  | Adjusted*  (n=24359161, events=3659300) | 2.45 (2.44 to 2.45) | <0.0001 |

**Figure S6. Sensitivity analysis time to event curves comparing acute hospitalisation.** Adjusted for age, sex, and baseline number of chronic diseases. Y-axis censored at 0.50.

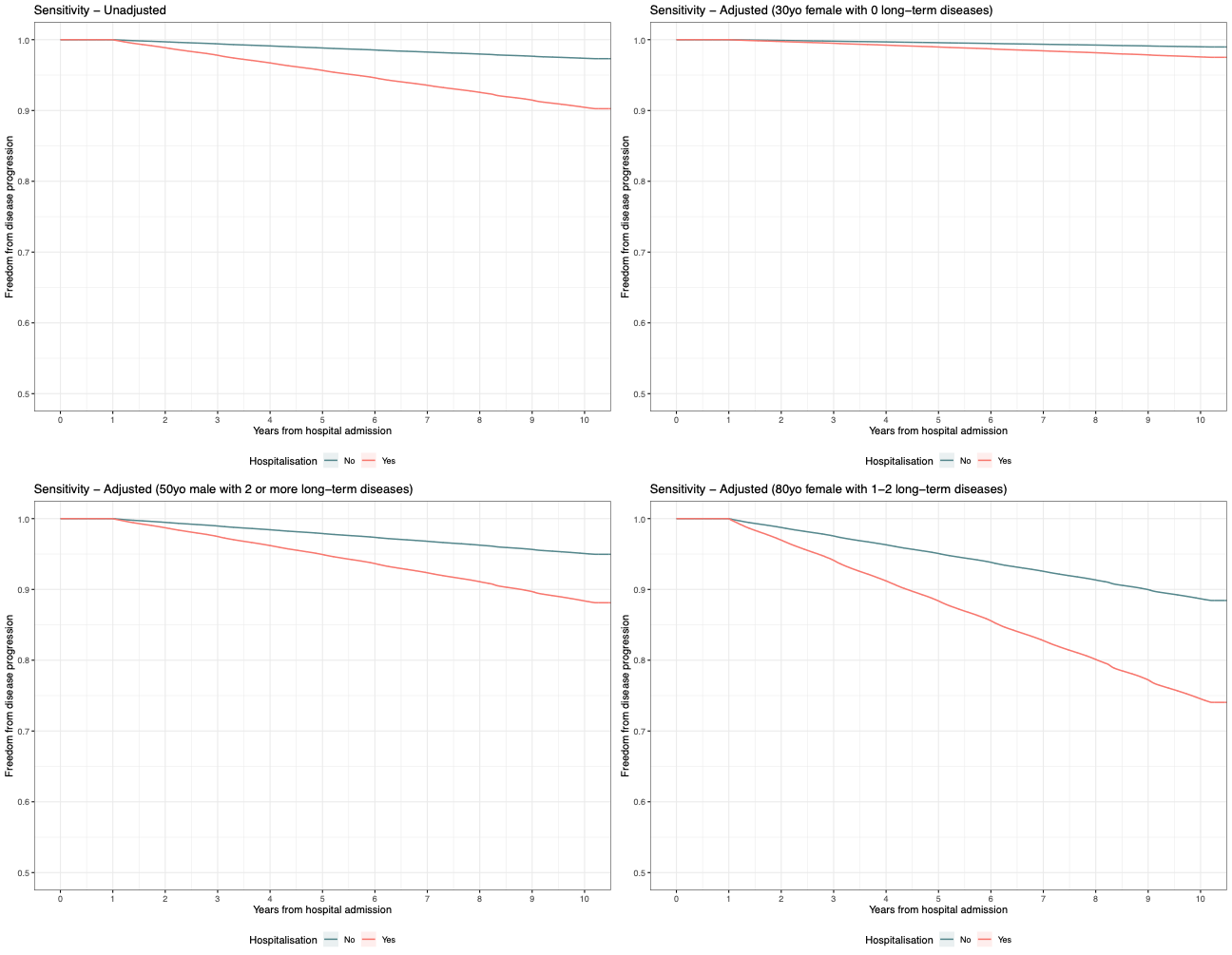

**Table S6. Secondary outcomes: total numbers of hospitalisations and multiple hospitalisation by IMD decile and ethnicity.** IMD: index of multiple deprivation, excluding unknown.

|  | **Acute hospitalisation** | |
| --- | --- | --- |
|  | Number  mean (SD) | Multiple  n (%) |
| All | 0.91 (2.6) | 3182865 (17.4) |
| *IMD* |  |  |
| 1 (most deprived) | 1.35 (3.8) | 380792 (24.1) |
| 2 | 1.07 (3.2) | 365139 (19.6) |
| 3 | 0.93 (2.8) | 345634 (17.4) |
| 4 | 0.88 (2.7) | 335972 (16.8) |
| 5 | 0.89 (2.5) | 323271 (17.0) |
| 6 | 0.84 (2.3) | 294297 (16.4) |
| 7 | 0.84 (2.2) | 303009 (16.6) |
| 8 | 0.80 (2.1) | 287777 (15.8) |
| 9 | 0.80 (2.1) | 272814 (15.9) |
| 10 (least deprived) | 0.74 (2.0) | 271396 (14.9) |
| 1-4 | 1.04 (3.1) | 1427537 (19.2) |
| 5-10 | 0.82 (2.2) | 1752564 (16.1) |
| *Ethnicity* |  |  |
| Asian | 0.64 (1.9) | 216234 (12.6) |
| Black | 0.88 (3.1) | 138109 (16.3) |
| Mixed | 0.68 (2.0) | 39924 (13.2) |
| Other | 0.19 (0.7) | 5075 (3.7) |
| Unknown | 0.13 (0.8) | 21865 (2.4) |
| White | 1.01 (2.8) | 2761658 (19.2) |

**Figure S7. Change in healthcare contact days comparing the year before and after acute hospitalisation.**

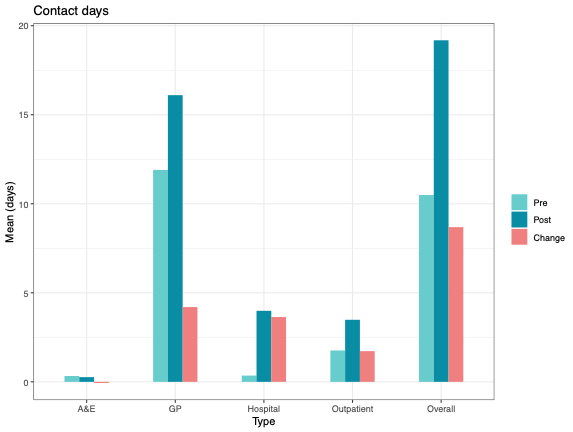

**Figure S8. Change in healthcare contact days comparing the year before and after acute hospitalisation comparing IMD deciles.** IMD: index of multiple deprivation.

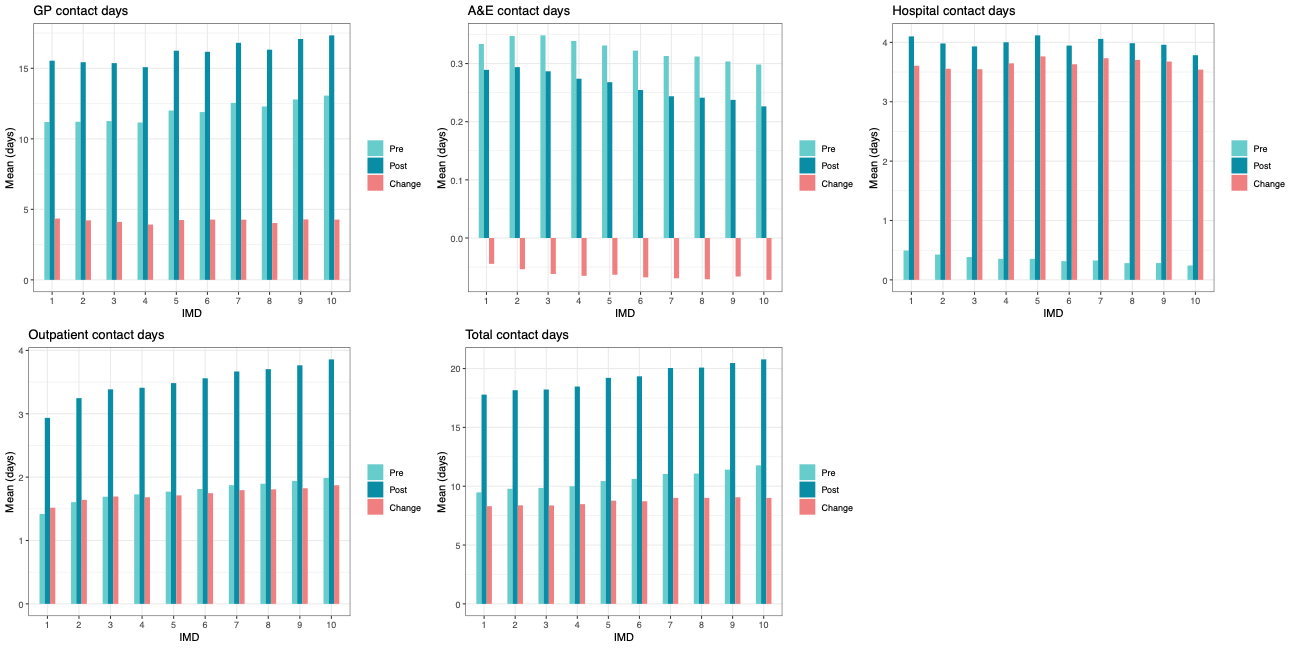

**Figure S9. Change in healthcare contact days comparing the year before and after acute hospitalisation comparing ethnic groups.**

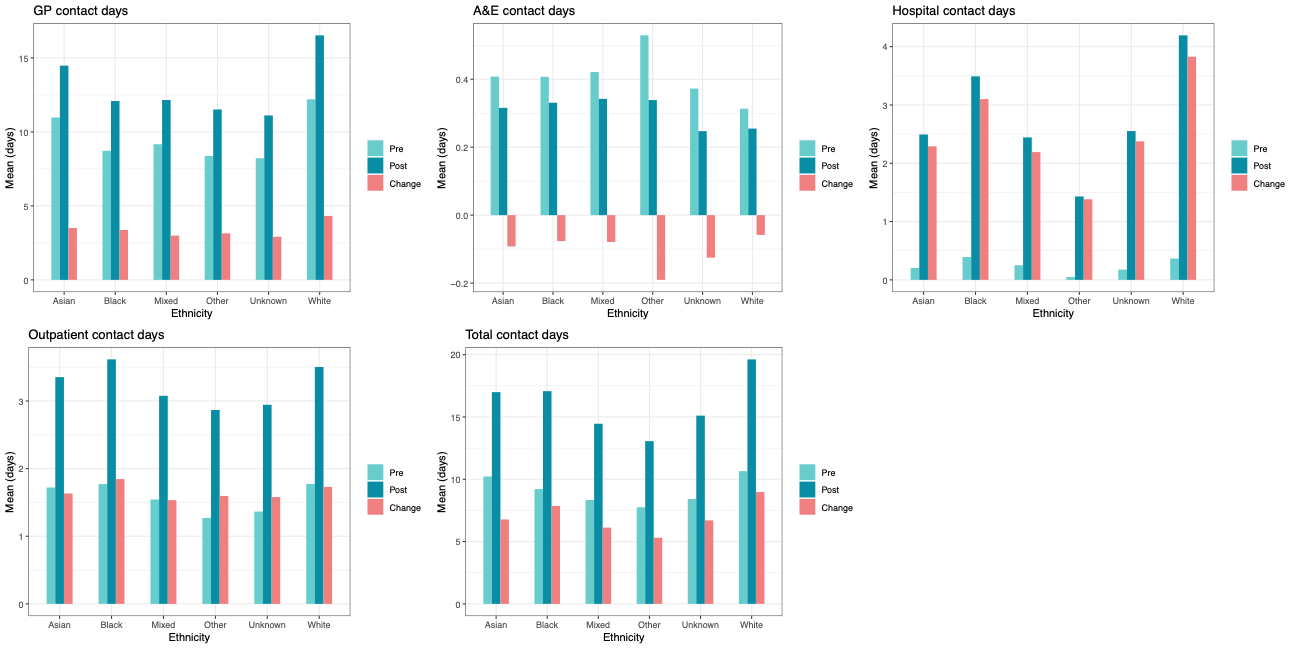

**Table S7. Age at which 50% of patients had experienced hospitalisation by IMD decile and ethnicity.** Analysis limited to preceding eight years where complete data was available. Other ethnicity excluded due to substantially fewer proportion of hospitalisations across the age range with <50% throughout.

|  | Proportion admitted at age (years) | | |
| --- | --- | --- | --- |
|  | 50% | 33% | 67% |
| *IMD* | | | |
| 1 (most deprived) | 70 | <50 | 80 |
| 2 | 74 | 56 | 82 |
| 3 | 76 | 61 | 83 |
| 4 | 77 | 64 | 84 |
| 5 | 78 | 66 | 84 |
| 6 | 78 | 68 | 85 |
| 7 | 79 | 69 | 85 |
| 8 | 79 | 70 | 85 |
| 9 | 80 | 71 | 86 |
| 10 (least deprived) | 80 | 72 | 86 |
| *Ethnicity* | | | |
| Asian | 78 | 63 | 86 |
| Black | 79 | 63 | 85 |
| Mixed | 80 | 69 | 88 |
| White | 77 | 64 | 84 |

We carried out an exploratory analysis to describe differences in age of first hospitalisation between IMD deciles and ethnicity groups considering differences in age distribution between IMD and ethnicity subgroups. Based on the age of all patients within the cohort at the end of follow up in 2021, we assessed the percentage of patients of each age who had experienced a hospital admission up to this date. We report the age at which 50% of people within each IMD decile and ethnic group had experienced hospitalisation.

**Figure S10. Proportion of patients who had hospital admission during the preceding eight years by age in 2021.** Panel A: Grouped by Index of Multiple Deprivation decile (IMD). Panel B: Grouped by major ethnicity category.

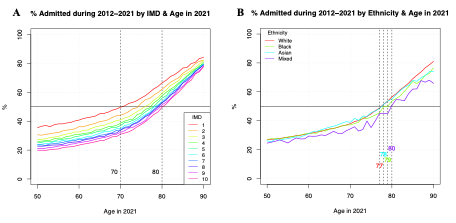

**The RECORD statement – checklist of items, extended from the STROBE statement, that should be reported in observational studies using routinely collected health data.**

|  | **Item No.** | **STROBE items** | **Location in manuscript where items are reported** | **RECORD items** | **Location in manuscript where items are reported** |
| --- | --- | --- | --- | --- | --- |
| **Title and abstract** | | | | | |
|  | 1 | (a) Indicate the study’s design with a commonly used term in the title or the abstract (b) Provide in the abstract an informative and balanced summary of what was done and what was found | Main text pages 1-2 | RECORD 1.1: The type of data used should be specified in the title or abstract. When possible, the name of the databases used should be included.  RECORD 1.2: If applicable, the geographic region and timeframe within which the study took place should be reported in the title or abstract.  RECORD 1.3: If linkage between databases was conducted for the study, this should be clearly stated in the title or abstract. | Main text pages 1-2  Main text pages 1-2  Main text pages 1-2 |
| **Introduction** | | | | | |
| Background rationale | 2 | Explain the scientific background and rationale for the investigation being reported | Main text page 4 |  |  |
| Objectives | 3 | State specific objectives, including any prespecified hypotheses | Main text page 4 |  |  |
| **Methods** | | | | | |
| Study Design | 4 | Present key elements of study design early in the paper | Main text page 5 |  |  |
| Setting | 5 | Describe the setting, locations, and relevant dates, including periods of recruitment, exposure, follow-up, and data collection | Main text page 5 |  |  |
| Participants | 6 | *(a) Cohort study* - Give the eligibility criteria, and the sources and methods of selection of participants. Describe methods of follow-up  *Case-control study* - Give the eligibility criteria, and the sources and methods of case ascertainment and control selection. Give the rationale for the choice of cases and controls  *Cross-sectional study* - Give the eligibility criteria, and the sources and methods of selection of participants  *(b) Cohort study* - For matched studies, give matching criteria and number of exposed and unexposed  *Case-control study* - For matched studies, give matching criteria and the number of controls per case | Main text page 5, Figure 1  N/A | RECORD 6.1: The methods of study population selection (such as codes or algorithms used to identify subjects) should be listed in detail. If this is not possible, an explanation should be provided.  RECORD 6.2: Any validation studies of the codes or algorithms used to select the population should be referenced. If validation was conducted for this study and not published elsewhere, detailed methods and results should be provided.  RECORD 6.3: If the study involved linkage of databases, consider use of a flow diagram or other graphical display to demonstrate the data linkage process, including the number of individuals with linked data at each stage. | Main text page 5  N/A  Main text page 5, Figure S1 |
| Variables | 7 | Clearly define all outcomes, exposures, predictors, potential confounders, and effect modifiers. Give diagnostic criteria, if applicable. | Main text pages 5-6 | RECORD 7.1: A complete list of codes and algorithms used to classify exposures, outcomes, confounders, and effect modifiers should be provided. If these cannot be reported, an explanation should be provided. | Supplement Table 1 |
| Data sources/ measurement | 8 | For each variable of interest, give sources of data and details of methods of assessment (measurement).  Describe comparability of assessment methods if there is more than one group | Main text page 5 |  |  |
| Bias | 9 | Describe any efforts to address potential sources of bias | Main text pages 5-6 |  |  |
| Study size | 10 | Explain how the study size was arrived at | Main text page 5 |  |  |
| Quantitative variables | 11 | Explain how quantitative variables were handled in the analyses. If applicable, describe which groupings were chosen, and why | Main text pages 5-6 |  |  |
| Statistical methods | 12 | (a) Describe all statistical methods, including those used to control for confounding  (b) Describe any methods used to examine subgroups and interactions  (c) Explain how missing data were addressed  (d) *Cohort study* - If applicable, explain how loss to follow-up was addressed  *Case-control study* - If applicable, explain how matching of cases and controls was addressed  *Cross-sectional study* - If applicable, describe analytical methods taking account of sampling strategy  (e) Describe any sensitivity analyses | Main text pages 5-6  Main text pages 5-6  Main text pages 5-6  N/A  Main text page 6 |  |  |
| Data access and cleaning methods |  | .. |  | RECORD 12.1: Authors should describe the extent to which the investigators had access to the database population used to create the study population.  RECORD 12.2: Authors should provide information on the data cleaning methods used in the study. | Main text page 12  Main text page 5 |
| Linkage |  | .. |  | RECORD 12.3: State whether the study included person-level, institutional-level, or other data linkage across two or more databases. The methods of linkage and methods of linkage quality evaluation should be provided. | Main text page 5 |
| **Results** | | | | | |
| Participants | 13 | (a) Report the numbers of individuals at each stage of the study (*e.g.*, numbers potentially eligible, examined for eligibility, confirmed eligible, included in the study, completing follow-up, and analysed)  (b) Give reasons for non-participation at each stage.  (c) Consider use of a flow diagram | Main text page 7, Figure S1 | RECORD 13.1: Describe in detail the selection of the persons included in the study (*i.e.,* study population selection) including filtering based on data quality, data availability and linkage. The selection of included persons can be described in the text and/or by means of the study flow diagram. | Main text page 5 |
| Descriptive data | 14 | (a) Give characteristics of study participants (*e.g.*, demographic, clinical, social) and information on exposures and potential confounders  (b) Indicate the number of participants with missing data for each variable of interest  (c) *Cohort study* - summarise follow-up time (*e.g.*, average and total amount) | Main text page 7, Table 1  Main text page 7, Tables 1-2, Supplement  Main text page 5 |  |  |
| Outcome data | 15 | *Cohort study* - Report numbers of outcome events or summary measures over time  *Case-control study* - Report numbers in each exposure category, or summary measures of exposure  *Cross-sectional study* - Report numbers of outcome events or summary measures | Main text pages 7-8, Table 2, Supplement |  |  |
| Main results | 16 | (a) Give unadjusted estimates and, if applicable, confounder-adjusted estimates and their precision (e.g., 95% confidence interval). Make clear which confounders were adjusted for and why they were included  (b) Report category boundaries when continuous variables were categorized  (c) If relevant, consider translating estimates of relative risk into absolute risk for a meaningful time period | Main text pages 7-8, Supplement  Main text pages 7-8, Supplement  N/A |  |  |
| Other analyses | 17 | Report other analyses done—e.g., analyses of subgroups and interactions, and sensitivity analyses | Main text page 8 |  |  |
| **Discussion** | | | | | |
| Key results | 18 | Summarise key results with reference to study objectives | Main text page 9 |  |  |
| Limitations | 19 | Discuss limitations of the study, taking into account sources of potential bias or imprecision. Discuss both direction and magnitude of any potential bias | Main text page 10 | RECORD 19.1: Discuss the implications of using data that were not created or collected to answer the specific research question(s). Include discussion of misclassification bias, unmeasured confounding, missing data, and changing eligibility over time, as they pertain to the study being reported. | Main text page 10 |
| Interpretation | 20 | Give a cautious overall interpretation of results considering objectives, limitations, multiplicity of analyses, results from similar studies, and other relevant evidence | Main text pages 9-11 |  |  |
| Generalisability | 21 | Discuss the generalisability (external validity) of the study results | Main text pages 9-11 |  |  |
| **Other Information** | | | | | |
| Funding | 22 | Give the source of funding and the role of the funders for the present study and, if applicable, for the original study on which the present article is based | Main text page 6 |  |  |
| Accessibility of protocol, raw data, and programming code |  |  |  | RECORD 22.1: Authors should provide information on how to access any supplemental information such as the study protocol, raw data, or programming code. | Main text page 12 |

*Reference: Benchimol EI, Smeeth L, Guttmann A, Harron K, Moher D, Petersen I, Sørensen HT, von Elm E, Langan SM, the RECORD Working Committee. The REporting of studies Conducted using Observational Routinely-collected health Data (RECORD) Statement. *PLoS Medicine* 2015; in press.

*Checklist is protected under Creative Commons Attribution ([CC BY](http://creativecommons.org/licenses/by/4.0/)) license.
